# Prevention of the dry season peak in child wasting in Chad: Evidence from a cluster randomised controlled trial of integrated livestock interventions

**DOI:** 10.64898/2026.04.07.26349927

**Authors:** Gwenaelle Luc, Modibo Keita, Brahima Diarra, Prospère Djekornonde, Fatima Abdelrazakh, Armelle Sacher, Bibata Wassonguema, Baguinebie Bazongo, Moursal Akoina, Mahamat Garba Issa, Moussa Abderamane, Cyprien Biaou, Tiphaine Seyvet, Abdelkader Abakar, Vincent Moutede, Camille Heylen, Matthew Bentley, Christine Jost, Helen Young, Mahamat Bechir, Mahamat Fayiz Abakar, Anastasia Marshak, Clair Null, Abdal Monium Osman

## Abstract

**Background:** Child acute malnutrition remains persistently above emergency thresholds in Chad’s Sahelian drylands, with a predictable, but rarely recognized, dry season peak linked to declining pasture and livestock productivity, reduced milk availability and heightened exposure to zoonotic infections. Humanitarian responses remain largely reactive and treatment-focused, with limited evidence on preventive strategies that address drivers embedded in local livelihood systems. We evaluated the effectiveness and return on investment (ROI) of an integrated livestock management intervention designed to prevent the dry-season peak of child acute malnutrition in pastoral and agro-pastoral communities in Chad.

**Methods:** We conducted a cluster-randomised controlled trial in Kanem and Barh-El-Gazel provinces, Chad. Seventy-six villages were randomised (1:1) to intervention or control. Eligible households had at least one child aged 6-59 months and access to milking livestock during the dry season. The intervention (December 2024-June 2025) combined livestock feed supplementation to sustain milk production near households during the dry season, household-level zoonotic risk mitigation, and nutrition counselling. Primary outcomes were the prevalence of global acute malnutrition (GAM) and severe acute malnutrition (SAM) at the dry-season peak (May 2025), assessed in a prespecified random subsample of 52 clusters. All 76 clusters were assessed post-peak (July 2025). Analyses followed an intention-to-treat approach using mixed-effects models. A societal ROI analysis was conducted over six months with projections to 24 months.

**Findings:** At the dry-season peak, 821 children 6-59 months from 521 households were assessed across 52 villages. GAM prevalence was 22.2% in intervention villages versus 47.4% in controls (adjusted OR 0.29 [95% CI 0.18-0.49]; p<0.001), and SAM prevalence was 4.4% versus 19.4% (adjusted OR 0.17 [0.08-0.37]; p<0.001). Intervention households had higher daily milk availability (+588 mL per household; p<0.001), and children consumed more milk (+102 mL per day; p=0.008). Odds of self-reported diarrhoeal disease and acute respiratory infection were substantially lower among children in intervention villages (aOR 0.21 [0.10-0.44] and 0.22 [0.11-0.46], respectively). Post-peak, women’s dietary diversity increased (aOR 3.68 [1.90-7.13]), alongside reduced workload, lower household food insecurity and distress livestock sales, improved livestock condition, and a benefit-cost ratio of 5.40 at six months, rising to 16.40 at 24 months.

**Interpretation:** Protecting livestock productivity and sustaining children’s access to milk while reducing zoonotic exposure during the pastoral lean season effectively prevents seasonal peaks of child acute malnutrition. This integrated anticipatory action and One Health livelihood-based approach offers a scalable, dignifying, high-return lifesaving preventive model for pastoral and agro-pastoral humanitarian settings.

## INTRODUCTION

Acute malnutrition remains one of the most immediate and life-threatening forms of undernutrition, affecting an estimated 42·8 million children under five globally in 2024 [1]. The burden is disproportionately concentrated in arid and semi-arid drylands, where recurrent shocks, precarious livelihoods, and pronounced seasonality drive persistently high prevalence of wasting and excess child mortality. Chad is among the most severely affected countries. In its Sahelian and Sahelo-Saharan zones, dominated by pastoral and agro-pastoral livelihoods, prevalence of global acute malnutrition (GAM) among children under five consistently exceeds the World Health Organisation (WHO) emergency thresholds [2]. These zones are persistently classified as Integrated Food Security Phase Classification for Acute Malnutrition (IPC AMN) Phase 3 (Crisis) or higher [3, 4], contributing substantially to preventable child deaths [5].

Humanitarian nutrition responses in Chad and similar dryland contexts have predominantly relied on reactive, treatment-centered approaches focused on treating malnutrition [6, 7]. These strategies, though they are lifesaving, have proven unable to break the cycle of persistent malnutrition. Anticipatory approaches remain marginal, accounting for less than 1% of global humanitarian financing [8], despite repeated calls to rebalance investments. This challenge is further compounded by an unprecedented contraction in humanitarian nutrition funding in 2025 [9]. In line with current humanitarian reset agendas^1^, there is growing pressure to prioritise cost-effective, scalable interventions that reduce future caseloads rather than perpetually expanding treatment capacity.

In dryland settings, acute malnutrition follows predictable seasonal patterns closely aligned with livelihood systems and environmental dynamics [10]. In Chad, as across much of the African Sahel, two seasonal peaks of child acute malnutrition occur each year [11–13]. The better-recognized peak arises toward the end of the rainy season, preceding the harvest and coinciding with the tail-end of the conventional hunger gap. By contrast, a primary, and often more severe, peak occurs at the end of the dry season, during the pastoral lean period. Despite its predictability and severity, this dry-season peak remains under-addressed in programme design and policy. Addressing this gap requires interventions that act on the structural drivers of acute malnutrition embedded within dryland livelihood systems as illustrated in the adapted framework for acute malnutrition in arid and semi-arid lands [14–16].

Livestock plays a central role in pastoral and agro-pastoral livelihoods, underpinning food security, nutrition, income, workload and time allocation, social relations, and cultural identity. In Chad, livestock support the livelihoods of more than 80% of rural households and represent the country’s most important agricultural subsector and largest non-oil economic activity [17]. Livestock can improve child nutrition outcomes through multiple, interconnected pathways [18], including direct provision of nutrient-dense foods [19], income generation that improves resilience capital [20], women’s empowerment [21, 22], and access to diverse diets. Milk, in particular, is a critical source of high-quality protein and essential micronutrients for young children, yet its availability is highly seasonal and declines sharply during the dry season^2^, coinciding with peak malnutrition risk. Evidence from African drylands suggests that sustaining livestock health and milk production close to households during this period can improve child diets and nutritional outcomes [23, 24]. At the same time, the pastoral lean season is characterized by heightened infection risk at the human–animal–environment interface^3^. Close proximity between children and livestock, reliance on unsafe water, and seasonal crowding around limited grazing and water points increase exposure to zoonotic and enteric pathogens associated with diarrhoeal disease, impaired nutrient absorption, and elevated mortality risk. [25–30]. These infection-related pathways frequently undermine nutritional gains achieved through food-based interventions alone. Integrating livestock management with measures to reduce environmental and zoonotic exposure therefore represents a potentially powerful, yet under-tested, prevention strategy [31].

Recent reflections informing the WHO Guideline on the prevention and management of wasting highlight a critical evidence gap: few rigorously evaluated interventions demonstrate meaningful reductions in acute malnutrition through preventive action [32]. Nutrition-sensitive programmes have shown improvements in intermediate outcomes such as child diet but limited effects on acute malnutrition itself. Experimental evidence is particularly scarce for integrated, multisectoral approaches that act upstream on predictable seasonal risks embedded in local livelihood systems, as well as for their cost-effectiveness.

To address this gap, we evaluated an integrated livelihood intervention designed as an anticipatory package implemented ahead of the pastoral dry-season peak of acute malnutrition. The intervention combined dry season livestock feed support to sustain milk availability near households, household level measures to reduce zoonotic and environmental infection risk, and targeted nutrition counselling to promote safe milk consumption and appropriate infant and young child feeding. We conducted a cluster-randomised controlled trial in pastoral and agro-pastoral communities in Kanem and Barh El-Gazel provinces, Chad, to test two hypotheses: first, that an integrated livestock-based intervention can prevent the dry season peak of child acute malnutrition; and second, that this anticipatory, livelihood-grounded approach generates substantial economic returns, assessed through a return-on-investment framework.

## METHODS

### Study design and setting

We conducted a parallel-group, cluster-randomized controlled trial (cRCT) in Kanem and Barh-El-Gazel provinces, Chad, a Sahelian dryland context characterized by pastoral and agro-pastoral livelihoods and persistent high burden of acute malnutrition that regularly exceeds the emergency threshold of 15% child wasting. Villages were used as the unit of randomization to reduce contamination between households and because key intervention pathways (livestock management, water practices, and behaviour change) operate at community level^4^.The interventions were implemented from December 2024 to June 2025, spanning the dry-season pastoral lean period, when the risk of acute malnutrition also peaks. To prioritize measurement during the period of dry season peak of acute malnutrition, primary outcomes were assessed at the dry season peak (May 2025) in a prespecified, random sample of 52 clusters (26 per arm - 521 households and 821 children surveyed) to estimate intervention effects^5^. A second, post-intervention assessment was conducted in July 2025 across all 76 randomised clusters (38 per arm - 1,243 households and 2,225 children surveyed) to assess sustained effects.

### Participants and eligibility

A baseline census conducted in 182 villages identified 6207 households, and 10529 children under 5. From this sampling frame, 76 villages were randomly selected for inclusion in the trial. Following selection, two villages became inaccessible due to emerging insecurity before enrolment and were replaced with randomly selected villages from the same baseline stratum to preserve balance and representativeness. Within each selected village, households were eligible if they met the following criteria: (1) had at least one child aged 6–53 months at enrolment (December 2024), ensuring age eligibility of 6–59 months at endline; (2) had access to at least one milking Tropical Livestock Unit (TLU)^6^ during the dry season; and (3) were identified as vulnerable through community-based targeting, based on community assessments of food insecurity, recent shocks, and socioeconomic vulnerability, using locally agreed criteria applied identically across the study arms. Written informed consent was obtained from caregivers; where literacy was limited, consent was documented through an approved witnessed procedure.

### Randomization and masking

Villages were randomized centrally in a stratified randomization scheme with 1:1 allocation to intervention or control arms (38 clusters per arm) (Figure 1). Stratification was conducted using key baseline indicators, including GAM and SAM prevalence, household food security (Food Consumption Score [FCS]), and livestock ownership (species composition and Tropical Livestock Units [TLUs]). Group comparability at baseline was assessed using the Household Hunger Scale (HHS), the reduced Coping Strategies Index (rCSI), primary income sources, cultivated land area, and diversity of community water sources. Given the nature of the intervention and village level allocation, blinding of participants and implementing teams was not feasible after assignment. To reduce measurement bias, enumerators responsible for anthropometry and household surveys were trained on standardized protocols and were not involved in intervention delivery. Measurement teams followed structured procedures with routine supervision and quality checks.

**Figure 1:**
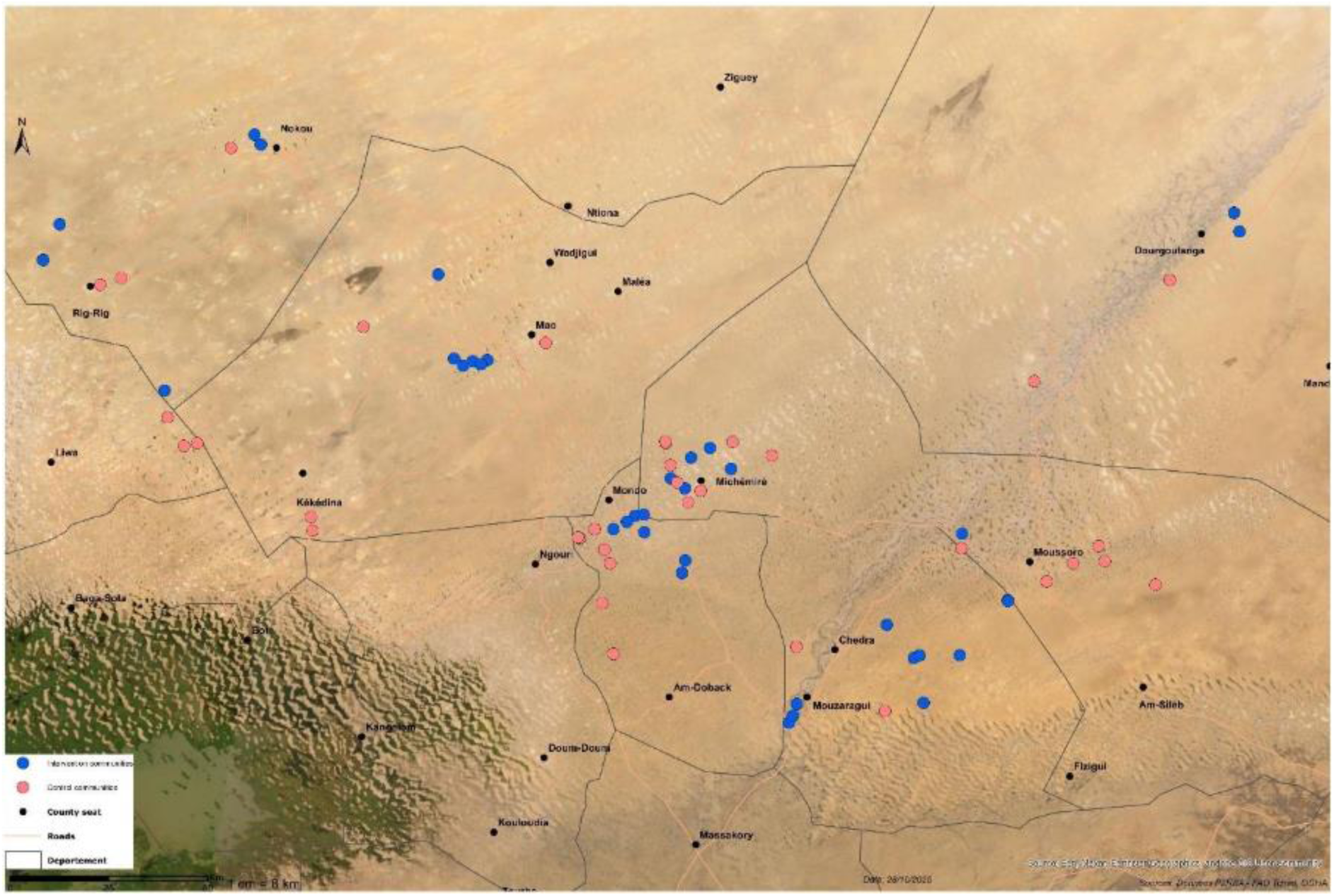
Spatial distribution of intervention and control villages.

### Intervention and control

The study adopted a formative mixed-methods approach combining quantitative and qualitative components to inform the intervention design. Although the formative findings are not presented in this paper, they were instrumental in identifying key livelihood-based pathways and refining the intervention components and parameters to improve nutrition outcomes in local pastoral and agro-pastoral communities.

The intervention targeted key dry-season pathways affecting livelihood production, exposure to enteric pathogens, and caregiving and feeding practices through three synergistic components delivered between December 2024 and May 2025. ^7^Figure 2 provides an overview of the interventions and implementation calendar.

**Figure 2:**
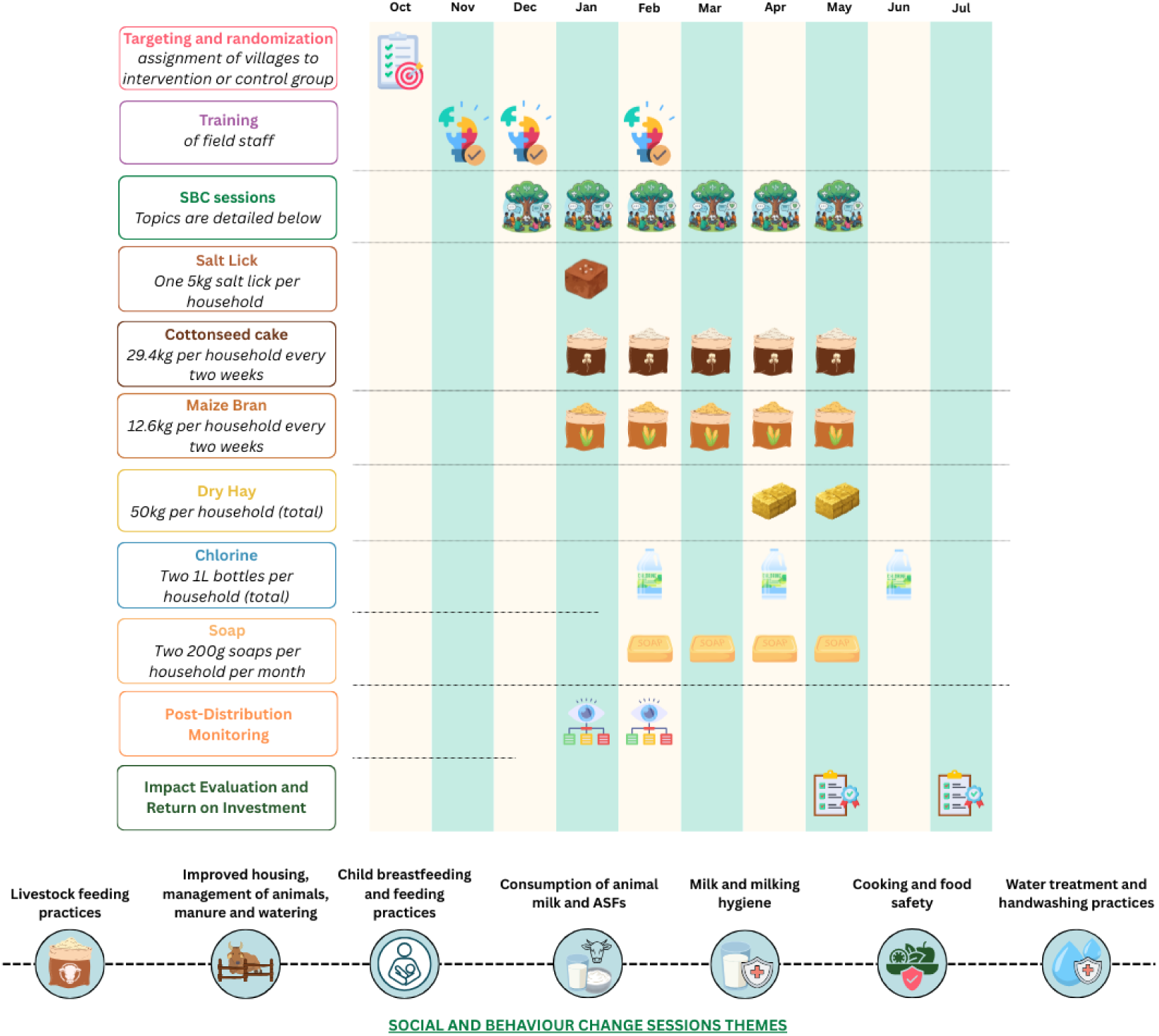
Detailed calendar of intervention.

First, livestock feed supplements and counselling were provided to eligible households to sustain milk production near homesteads during the dry season. Supplementary feed was distributed biweekly at the village level and accompanied by counselling on optimal livestock feeding and milking practices to enhance productivity and ensure safe household milk consumption. Households with access to at least one milking TLU received biweekly distributions of supplementary livestock feed. The daily ration per milking TLU comprised 0.9 kg cottonseed cake and 2.1 kg maize bran, supplemented by a 5 kg mineral salt lick per household for the entire intervention period. Additionally, 50 kg of dry hay was provided per household in April to mitigate peak pasture scarcity.

Second, zoonotic and enteric risk mitigation was implemented through the provision of chlorine (i.e., bleach) for drinking water treatment and soap, supported by monthly community level behaviour change counselling. Counselling focused on prioritization of safer sources, water treatment and safe storage, hand hygiene, safe milk handling, safe food preparation and consumption, and separation of human and animal water sources. Third, nutrition counselling promoted safe milk consumption and age-appropriate infant and young child feeding (IYCF) practices, including exclusive breastfeeding and complementary feeding, alongside dietary practices for pregnant and breastfeeding women. Hand washing, breastfeeding and child feeding tools and messages followed the national curriculum, with additional modules developed. Counselling was delivered every two weeks at village level in intervention villages only, and was open to all community members from intervention villages.

Counseling were delivered through a structured series of six-monthly group sessions, each lasting approximately 45 minutes, complemented by bi-weekly reinforcement sessions (10–15 minutes) conducted during inputs distributions, for a total of 12 reminders sessions. Sessions were delivered at community level and were open to all members of the intervention communities, including both targeted households and non-beneficiaries. Each monthly session introduced a new thematic topic, while reinforcement sessions focused on reviewing and reinforcing key messages across all topics. All counselling and distributions were scheduled flexibly to align with women’s availability and preferences, ensuring accessibility and minimizing disruption to their daily responsibilities.

Villages in the control cluster continued to receive routine services available in the area including routine livestock vaccination and deworming campaigns, where implemented. To ensure ethical equipoise between study arms, control households received livestock feed after completion of study follow-up.

### Outcomes

Primary outcomes were assessed among children aged 6–59 months at the dry-season peak (May 2025) and consisted of:

- Global acute malnutrition (GAM): WHZ < −2 and/or MUACZ < −2 and/or bilateral pitting oedema
- Severe acute malnutrition (SAM): WHZ < −3 and/or MUACZ < −3 and/or bilateral pitting oedema All components were assessed for all children using the same measurements and procedures. We also reported WHZ-only, MUACZ-only and Oedema for interpretability

### Secondary outcomes were prespecified to reflect hypothesized pathways of impact and included

household milk availability and child milk consumption; child morbidity (two-week caregiver recall) including reported diarrhoea, acute respiratory and fever symptoms within the past two weeks, fever; women’s dietary diversity (Minimum Dietary Diversity for Women [MDD-W]); women’s workload and perceived physical exhaustion; household food security (Reduced coping strategy index [rCSI]; months of Adequate Household Food Provisioning [MAHFP]); herd health and productivity (body condition score, distress sales, births, morbidity); social cohesion and sharing practices (sharing of inputs, milk, childcare support); and household drinking water quality (microbiological testing for *E. coli,* fecal coliforms and total coliforms).

### Sample size

The trial was powered to detect a minimum absolute reduction of 5 percentage points in GAM prevalence between intervention and control arms with 80% power at a two-sided significance level of 0.05. Sample size calculations assumed a baseline GAM prevalence of 15%, based on routine surveillance data, and an intra-cluster correlation coefficient (ICC) of 0.05 to account for village-level clustering. Under these assumptions, 38 clusters per arm with 20 eligible households per cluster (1,520 households in total) were required.

### Data collection procedures

Child anthropometry followed WHO standardized procedures (age, sex, weight, length/height, MUAC, oedema). Enumerators received structured training and standardization for measurement with routine supervision and repeat measurements for quality assurance. Household questionnaires captured demographics characteristics, livestock production, milk consumption, child morbidity, food security, women’s diet and workload, and social cohesion. Drinking water samples were collected and analyzed for microbiological contamination (i.e., E. coli, fecal coliforms, and total coliforms) using the US EPA membrane filtration method ISO 9308-1:2014 and standard laboratory protocols. Adherence to household water treatment with bleach was assessed through measurement of free residual chlorine (FRC) in household stored drinking water during the May 2025 survey. Drinking water samples were collected at the household level and analysed on-site for FRC, PH, and turbidity using a HANNA chlorimeter, pH test strips, and turbidimeter, respectively, following standard field protocols.

**Figure 3:**
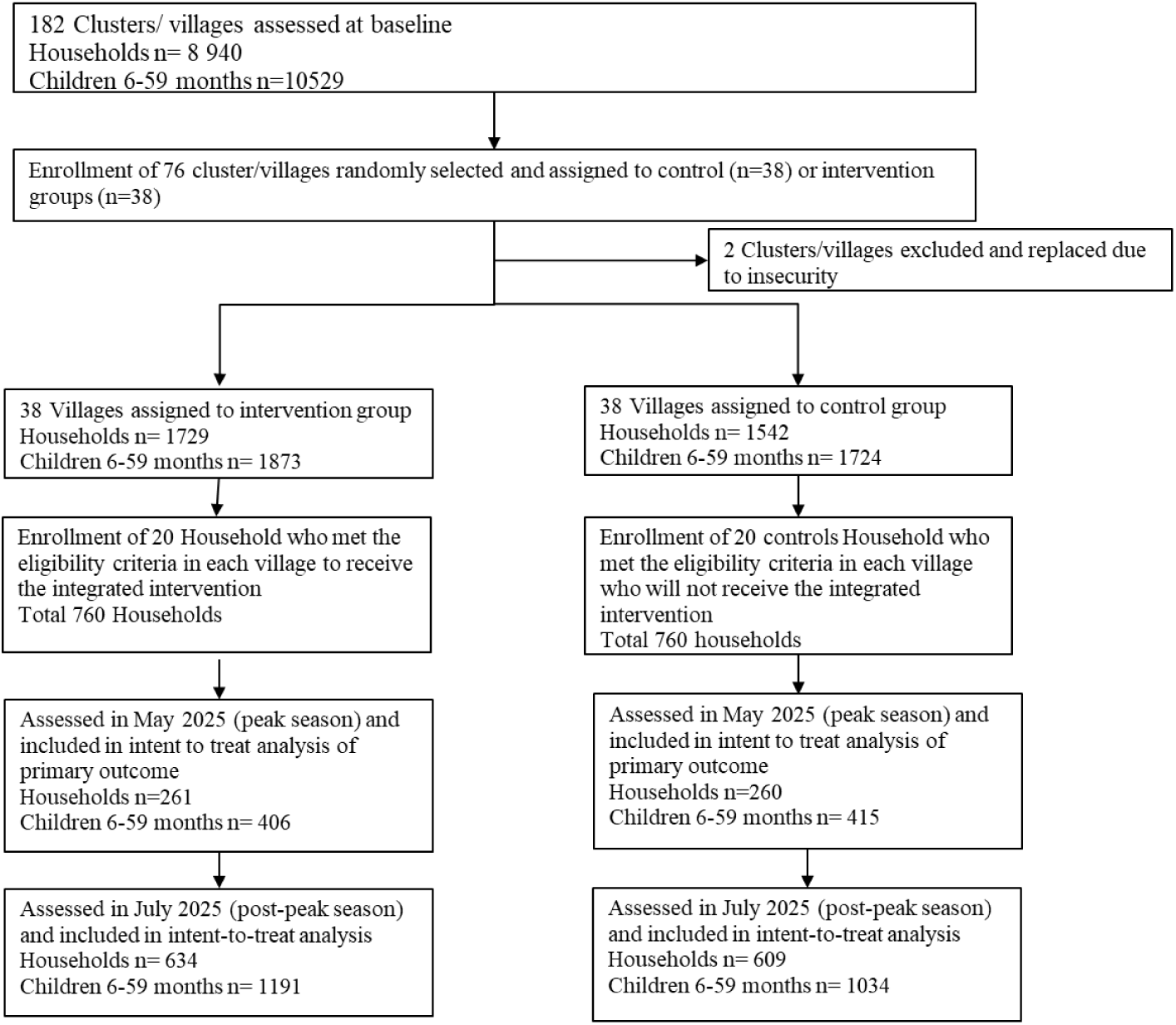
Consort Diagram Flow.

### Statistical analysis

Analyses followed an intention-to-treat principle, with children and households analyzed according to the randomized allocation of their village. Binary outcomes were analysed using generalized linear mixed models with a random intercept at the village level to account for clustering inherent to the trial design. Robust standard errors were estimated using appropriate sandwich estimators for cluster-randomized trials.

Baseline covariates were examined a priori to assess balance across study arms. These included household primary reliance on livestock for income, ethnic composition, prevalence of acute malnutrition, household food insecurity (rCSI, HHS, FCS), child morbidity symptoms, child dietary diversity, exclusive breastfeeding, median livestock ownership and main household water sources.

Similarly, at endline we assessed a prespecified set of child-, household-, and community-level characteristics, including child age and sex, mean number of children per household, household reliance on livestock as the main income source, household and community receipt of external assistance, mean NDVI within a 5-km buffer around each community (January–June 2025), and distance to the nearest health center.

Adjusted models included all the above covariates regardless of statistical significance.

Continuous secondary outcomes were analyzed using mixed-effects or generalized regression models aligned with their distributions. Anthropometric data quality was rigorously assessed through WHO flagging criteria, digit preference analysis, distributional checks, and dispersion diagnostics.

Prespecified subgroup analyses explored effect heterogeneity by child sex and age group. All analyses were conducted in R using fully reproducible workflows.

### Missing data

Analyses were performed on an available case basis for each outcome. The extent, distribution, and patterns of missing data were examined by study arm. No imputation was undertaken for missing outcomes. Missing data for primary outcomes was negligible (<1%) at May assessment but increased at the July follow-up (18.2% of the expected sample).

### Harm and adverse events

Field teams were trained to actively identify children presented with severe wasting or clinical illness and to refer them to routine health services in accordance with national protocols. Livestock requiring veterinary attention were similarly referred to the veterinary services. Adverse events potentially related to intervention components were monitored systematically and documented through supervisory and reporting systems throughout implementation. No adverse events attributable to the intervention were reported.

### Ethics and trial registration

The study protocol was approved by the Chad National Bioethics Committee and the Tufts University Institutional Review Board (IRB). The trial was registered with the Pan African Clinical Trials Registry (PACTR202504882709268).

### Societal Return on Investment (ROI) analysis

A societal benefit–cost analysis was conducted over the primary six-month intervention horizon (December 2024–May 2025), with modeled projections to 24 months. Future benefits were projected using conservative model-based extrapolation grounded in observed short-term trial effects and published epidemiological parameters. Costs included programme delivery (inputs, logistics, personnel, training) and household costs where applicable (e.g., time/opportunity costs). Research-related costs were not included. Benefits included monetized health and nutrition gains (e.g., reduced morbidity and treatment costs; modeled mortality risk reduction where applicable), and livestock and resilience-related gains (e.g., avoided distress sales, improved livestock condition and productivity), using observed trial effects, local market prices, and published parameters. Sensitivity analyses assessed robustness to key assumptions (detailed in appendix 1).

## RESULTS

### Baseline Characteristics and Comparability

Baseline characteristics were generally balanced between intervention and control villages (Table 1). Child anthropometrics indicators, including the prevalence of GAM and SAM, did not differ significantly between groups at baseline (p > 0.05). Baseline child morbidity symptoms and child dietary diversity were also similar across arms (p > 0.05). Household food security indicators, child morbidity symptoms, and livestock ownership patterns were broadly comparable across study arms, although small but statistically insignificant variations were observed. Village level ethnic composition also differed significantly between study arms.

**Table 1.** Baseline Characteristics comparability.

| Indicator | Intervention villages | Control villages | p-value | Odds ratio [95% CI] |
| --- | --- | --- | --- | --- |
| Child nutrition indicators aged 6-59 months |  |  |  |  |
| GAM prevalence (WHZ<-2) | 14.7% [13.2; 16.4] (n = 1,873) | 13.1% [11.5; 14.7] (n = 1,724) | 0.1578 | 1.15 [0.94; 1.39] |
| GAM prevalence (WHZ and/or MUACZ<-2) | 22.8% [20.9; 24.7] (n = 1,874) | 20.4% [18.6; 22.4] (n = 1,725) | 0.0834 | 1.15 [0.98; 1.35] |
| SAM prevalence (WHZ<-3) | 3.8 [3.1 ; 4.8] (n = 1873) | 2.7 [2.1 ; 3.6] (n = 1724) | 0.062 | 1.43 [0.98 ; 2.07] |
| SAM prevalence (WHZ and/or MUACZ <-3 and/or oedema) | 5 [4.1 ; 6.1] (n = 1871) | 4.3 [3.4 ; 5.4] (n = 1721) | 0.341 | 1.16 [0.85 ; 1.59] |
| Mean MUAC (mm) | 141.9 [141.3; 142.4] (n = 1,896) | 141.8 [141.3; 142.4] (n = 1,741) | 0.9304 | 0.83 [0.42; 1.63] |
| Oedema prevalence | 0.3% [0.1; 0.7] (n = 1,896) | 0.3% [0.1; 0.7] (n = 1,740) | 0.8732 | 1.10 [0.28; 4.57] |
| Children 0–5 months predominantly breastfed (water + breastmilk) | 38.6% [31.1; 46.8] (n = 145) | 38.7% [30.2; 48.1] (n = 111) | 0.9847 | 1.00 [0.58; 1.71] |
| Mean child dietary diversity score (IDDS) | 1.4 [1.3; 1.5] (n = 603) | 1.5 [1.4; 1.6] (n = 574) | 0.2193 | 1.24 [0.85; 1.80] |
| Children with low dietary diversity | 83.4% [79.7; 86.5] (n = 458) | 87.0% [83.6; 89.8] (n = 463) | 0.1203 | 0.75 [0.51; 1.10] |
| Children with diarrhea (past two weeks) | 30.9% [27.6; 34.5] (n = 676) | 31.8% [28.4; 35.5] (n = 660) | 0.7229 | 1.04 [0.82; 1.32] |
| Children with fever (past two weeks) | 56.4% [52.6; 60.1] (n = 676) | 56.1% [52.2; 59.8] (n = 660) | 0.9120 | 0.99 [0.79; 1.23] |
| Children with cough (past two weeks) | 49.6% [45.8; 53.3] (n = 676) | 53.9% [50.1; 57.7] (n = 660) | 0.1091 | 1.19 [0.96; 1.49] |
| Household food security indicators |  |  |  |  |
| Mean Food Consumption Score (FCS) | 41.3 [40.7; 41.8] (n = 1,729) | 42.1 [41.5; 42.6] (n = 1,542) | 0.0440 | 0.94 [0.78; 1.14] |
| Households with poor FCS | 12.1% [10.7; 13.8] (n = 1,729) | 11.4% [9.9; 13.1] (n = 1,542) | 0.5173 | 1.07 [0.86; 1.34] |
| Households with Household Hunger Score moderate or severe | 22 [20.1; 24.1] (n = 1729) | 25 [22.9; 27.2] (n = 1542) | 0.04812 | 0.85 [0.72; 1] |
| Households with rCSI Phase 3 | 3.9% [3.1; 4.9] (n = 1,729) | 4.5% [3.6; 5.7] (n = 1,542) | 0.3438 | 0.85 [0.59; 1.21] |
| Livelihoods indicators |  |  |  |  |
| Households with livestock as main income source | 19.8% [18.0; 21.8] (n = 1,729) | 29.2% [27.0; 31.6] (n = 1,542) | <0.001 | 0.60 [0.51; 0.71] |
| Median number of small ruminants owned (Q1;Q3) | 10.7 [10.0; 11.3] (n = 1,729) | 13.0 [12.3; 13.6] (n = 1,542) | 0.05937(based on the mean) | Mean difference -2.3 [-4.6; 0.1] |
| Median number of large ruminants owned (Q1;Q3) | 3.7 [3.3; 4.1] (n = 1,729) | 4.0 [3.6; 4.4] (n = 1,542) | 0.76658 (based on the mean) | Mean difference -0.2 [-1.9; 1.4], |
| Households using borehole as main water source | 72.4% [70.3; 74.5] (n = 1,729) | 72.0% [69.7; 74.2] (n = 1,542) | 0.7854 | 1.02 [0.87; 1.19] |
| Village level main ethnic group Composition |  |  |  |  |
| Yorda, Gourane, Yiria (plus “Other”) | 15.4 [13.8; 17.2] (n = 1,729) | 1.8 [1.2; 2.6] (n = 1,542) | 0,00000 | 10.2 [6.82; 15.25] |
| Kanembou (dominant) | 26.5 [24.5; 28.6] | 35.0 [32.6; 37.4] | 0,00000 | 0.67 [0.58; 0.78] |
| Boudoula, Yiria, Gourane (mixed) | 3.1 [2.4; 4.1] | 6.4 [5.2; 7.7] | 0,00002 | 0.48 [0.34; 0.67] |
| Gourane (dominant), Ankorda | 55.0 [52.6; 57.3] | 56.9 [54.5; 59.4] | 0,26559 | 0.92 [0.80; 1.06] |

These baseline imbalances were prespecified and accounted for in adjusted analyses.

### Endline characteristics and external assistance

At endline child age, sex distribution, household size, reliance on livestock as the primary income source, vegetation conditions (NDVI), and distance to health facilities were comparable (Table 2).

**Table 2.** Endline characteristics comparability.

| Variable | Survey round | Intervention group | Control group | p-value | Effect size / OR [95% CI] |
| --- | --- | --- | --- | --- | --- |
| Child characteristics |  |  |  |  |  |
| Girls (%) | May | 49.0 [44.2–53.9] (n=406) | 49.9 [45.1–54.7] (n=417) | 0.804 | OR 1.04 [0.78–1.37] |
|  | July | 47.2 [44.1–50.4] (n=978) | 51.1 [47.8–54.4] (n=877) | 0.085 | OR 0.85 [0.71–1.02] |
| Age (months), mean | May | 30.5 [29.0–32.1] (n=406) | 31.4 [29.7–33.1] (n=417) | 0.479 | $\Delta$ 0.82 [–1.46–3.11] |
| | July | 32.5 [31.6–33.4] (n=995) | 33.5 [32.5–34.5] (n=886) | 0.262 | $\Delta$ –1.0 [–2.8–0.8] |
| Household composition |  |  |  |  |  |
| Children per household, mean | May | 1.6 [1.5–1.8] (n=261) | 1.6 [1.4–1.8] (n=260) | 0.991 | $\Delta$ 0.00 [–0.24–0.24] |
| | July | 1.5 [1.4–1.5] (n=632) | 1.5 [1.4–1.5] (n=609) | 0.909 | $\Delta$ 0.00 [–0.10–0.10] |
| Livelihoods |  |  |  |  |  |
| Livestock as main income source (%) | July | 83.4 [80.3–86.1] (n=634) | 83.3 [80.1–86.0] (n=609) | 0.929 | OR 1.01 [0.74–1.38] |
| Programme exposure and assistance |  |  |  |  |  |
| Child enrolled in acute malnutrition treatment (%) | July | 4.8 [3.6–6.4] (n=975) | 6.4 [5.0–8.2] (n=875) | 0.462 | OR 0.74 [0.33–1.65] |
| Household received cash in past 6 months (%) | July | 10.0 [7.9–12.6] (n=632) | 6.4 [4.7–8.7] (n=609) | 0.680 | OR 1.40 [0.29–6.79] |
| Household received agricultural inputs in past 12 months (non-study) (%) | July | 15.2 [12.6–18.2] (n=632) | 4.1 [2.8–6.0] (n=609) | 0.091 | OR 3.70 [0.81–16.84] |
| Household received food assistance in the past 6 months (%) _ Note the distribution took place in July 2025 | July | 20.9 [17.9–24.2] (n=632) | 10.5 [8.3–13.2] (n=609) | <0.001 | OR 2.25 [1.61–3.16] |
| Village-level context |  |  |  |  |  |
| NDVI (Jan–Jun 2025), mean | — | 0.17 [0.17–0.17] (n=26) | 0.17 [0.17–0.17] (n=26) | 0.180 | $\Delta$ 0.00 [0.00–0.00] |
| Distance to nearest health facility (km), mean | — | 5.7 [3.7–7.7] (n=26) | 4.6 [2.7–6.5] (n=26) | 0.410 | Δ 1.1 [–1.6–3.8] |

Receipt of food assistance differed between arms, a higher proportion of intervention households reporting receipt of food assistance the preceding six months. As this assistance was delivered after the May peak assessment, it was included as an adjustment variable only in analyses of July outcomes.

### Intervention adherence

Adherence to the integrated livestock management intervention was high across promoted behaviors and practices (table 3). Intervention households kept a greater number of lactating TLU near the homestead during the dry season compared to controls. The median number of lactating TLUs kept near the household during the dry season was 0.9 in the intervention group versus 0.3 in control (p = 0.0029).

**Table 3:** Adherence to promoted livestock management and zoonotic risk prevention practices.

| Indicator | Intervention group | Control group | Effect / p-value | Notes |
| --- | --- | --- | --- | --- |
| Median number of lactating TLU kept near the household during the dry season | 0.9 (n=634) | 0.3 (n= 609) |  | This indicates adherence among intervention households to the promoted practice of keeping one TLU close to the homestead during the dry season. |
| Average number of lactating TLU kept near the household during the dry season | 1.1 [1.1; 1.2] (n = 634) | 0.7 [0.6; 0.8] (n = 609) | 0.4 [0.1; 0.7] (p_value = 0.0029) |  |
| Households reporting treating water with bleach | 69.1 % [63.2; 74.4] (n = 262) | <1 % [0; 1.8] (n = 258) | $p < 0.001$ | Indicates uptake of water-treatment behavior among intervention households. |
| Free residual chlorine (FRC, mg/L) measured in household water measured in May 2025 | 0.4 [0.2 – 0.5] (n = 262) | 0.1 [0.1 – 0.1] (n = 258) | Mean_diff : 0.3 [0.1; 0.5] p_value = 0.00008 | Confirms effective chlorination in intervention sites. *Note:* Some boreholes were treated at the source by other actors across both groups, which may explain detectable FRC levels in control groups. |
| Turbidity measured in household water – May 2025 | 6.7 [4.7 – 8.7] (n = 262) | 17.5 [0 – 40.3] (n = 258) | Mean_diff : -10.8 [-33.7; 12.1] p_value = 0.350 | Lower turbidity in intervention sites suggests adherence to messages on prioritizing safe water sources, reflecting a shift toward reliance on fewer but more protected water points |
| Households observed with handwashing station and soap – May 2025 | 75.4 % [71.8; 78.6] (n = 629) | 47.4 % [43.4; 51.3] (n = 606) | OR=3.4 [1.77; 6.52] (p_value = 0.00023) | Observation-based measure of hygiene adherence by enumerators |

Household level uptake of water treatment practices was substantially higher in intervention villages. Among households assessed for drinking water quality, 69.1% of intervention households reported treating drinking water with chlorine bleach, compared to none in the control group (p < 0.001). Objective measures confirmed higher free residual chlorine concentrations in stored household water in intervention households (mean difference 0.3 mg/L; p = 0.00008).

Observed hygiene practices also differed between groups. A higher proportion (75.4%) of intervention households had handwashing stations equipped with soap at the time of survey, compared to 47.4% among controls (p = 0.00023). Turbidity levels in household water were lower in intervention villages; but this difference was not statistically significant.

### Primary Outcomes at the Dry Season Peak (May 2025)

The prevalence of both GAM and SAM among children aged 6–59 months was substantially lower in intervention village than control (Table 4, both p < 0·001).

**Table 4:** Primary outcome in May 2025 – Peak dry season.

| Indicator | Intervention group (%) | Control group (%) | Odds ratio (unadjusted) [95% CI] | p-value | Odds ratio (adjusted) [95% CI] | p-value |
| --- | --- | --- | --- | --- | --- | --- |
| Overall |  |  |  |  |  |  |
| GAM (WHZ and/or MUACZ $< -2$ or oedema) | 22.2 | 47.4 | 0.33 [0.19–0.55] | $<0.001$ | 0.29 [0.18–0.49] | $<0.001$ |
| GAM (WHZ $< -2$) | 15.1 | 27.5 | 0.47 [0.29–0.74] | 0.001 | 0.43 [0.27–0.68] | $<0.001$ |
| GAM (MUACZ $< -2$) | 12.8 | 41.0 | 0.22 [0.12–0.39] | $<0.001$ | 0.19 [0.10–0.35] | $<0.001$ |
| SAM (WHZ and/or MUACZ $< -3$ or oedema) | 4.4 | 19.4 | 0.20 [0.09–0.41] | $<0.001$ | 0.17 [0.08–0.37] | $<0.001$ |
| SAM (WHZ $< -3$) | 3.2 | 7.5 | 0.41 [0.18–0.89] | 0.025 | 0.39 [0.18–0.86] | 0.020 |
| SAM (MUACZ $< -3$) | 1.2 | 15.1 | 0.07 [0.02–0.22] | $<0.001$ | 0.06 [0.02–0.19] | $<0.001$ |
| Oedema | 0.2 | 0.5 | 1.95 [0.18–20.86] | 0.580 | — | — |
| By Sex of the child |  |  |  |  |  |  |
| GAM (WHZ/MUACZ $< -2$) – Girls | 21.1 | 40.4 | 0.40 [0.20–0.81] | 0.010 | 0.38 [0.19–0.77] | 0.007 |
| GAM (WHZ/MUACZ $< -2$) – Boys | 23.3 | 54.3 | 0.26 [0.15–0.46] | $<0.001$ | 0.23 [0.13–0.40] | $<0.001$ |
| SAM (WHZ/MUACZ < -3 or oedema) – Girls | 5.5 | 13.5 | 0.39 [0.17–0.88] | 0.024 | 0.37 [0.16–0.86] | 0.020 |
| SAM (WHZ/MUACZ < -3 or oedema) – Boys | 3.4 | 25.4 | 0.10 [0.04–0.29] | <0.001 | 0.09 [0.03–0.26] | <0.001 |
| By Age group of children |  |  |  |  |  |  |
| GAM (WHZ/MUACZ < -2) – 6–23 months | 13.2 | 37.0 | 0.27 [0.12–0.62] | 0.002 | 0.25 [0.10–0.63] | 0.003 |
| GAM (WHZ/MUACZ < -2) – 24–59 months | 27.2 | 52.3 | 0.35 [0.20–0.60] | <0.001 | 0.30 [0.18–0.50] | <0.001 |
| SAM (WHZ/MUACZ < -3 or oedema) – 6–23 months | 4.2 | 9.0 | 0.44 [0.15–1.28] | 0.132 | 0.48 [0.15–1.57] | 0.224 |
| SAM (WHZ/MUACZ < -3 or oedema) – 24–59 months | 4.6 | 24.4 | 0.15 [0.06–0.36] | <0.001 | 0.12 [0.05–0.27] | <0.001 |

The prevalence of GAM among children aged 6-59 months was 22.2% in intervention villages compared with 47.4% in control villages, corresponding to a 25.2 percentage-point absolute reduction (table 4). After adjustment for clustering and baseline imbalances, children in intervention villages had 71% lower odds of GAM than those in the control group (p < 0.001).

SAM prevalence was 4.4% in intervention villages and 19.4% in controls, an absolute reduction of 15.0 percentage-points, The adjusted odds of SAM were 83% lower in intervention villages (p < 0.001).

Results were consistent across anthropometric definitions using WHZ-only and MUACZ-only criteria (all p ≤ 0·025). Oedema prevalence was low in both groups, and the difference between intervention and control villages was not statistically significant (0·2% vs 0·5% p = 0·58); estimates were imprecise because of small numbers.

### Subgroup analyses

Reductions in GAM were observed across child sex and age groups (Table 4), including among girls (p = 0·007) and boys (p < 0·001), and among children aged 6–23 months (p = 0·003) and 24–59 months (p < 0·001). Reductions in SAM were observed among boys (p < 0·001) and children aged 24–59 months (p < 0·001), but not among children aged 6–23 months (p = 0·224).

### Primary Outcomes After the Dry Season peak (July 2025)

Across all 76 villages, the prevalence of global acute malnutrition (GAM) remained significantly lower in intervention villages than in control villages after the dry season peak. GAM prevalence was 19·0% (95% CI 16·7–21·4; n = 1060) in intervention villages compared with 25·0% (22·3–28·0; n = 867) in control villages. After adjustment, children in intervention villages had 31% lower odds of GAM than those in control villages (adjusted OR 0·69; p = 0·039). Effect estimates were attenuated compared with those observed at the dry season peak.

### Secondary Outcomes

Secondary outcomes showed consistent improvements across livestock production, child health, women’s diets, and household resilience (Table 5 and Figure 4- Forest plot).

**Figure 4:**
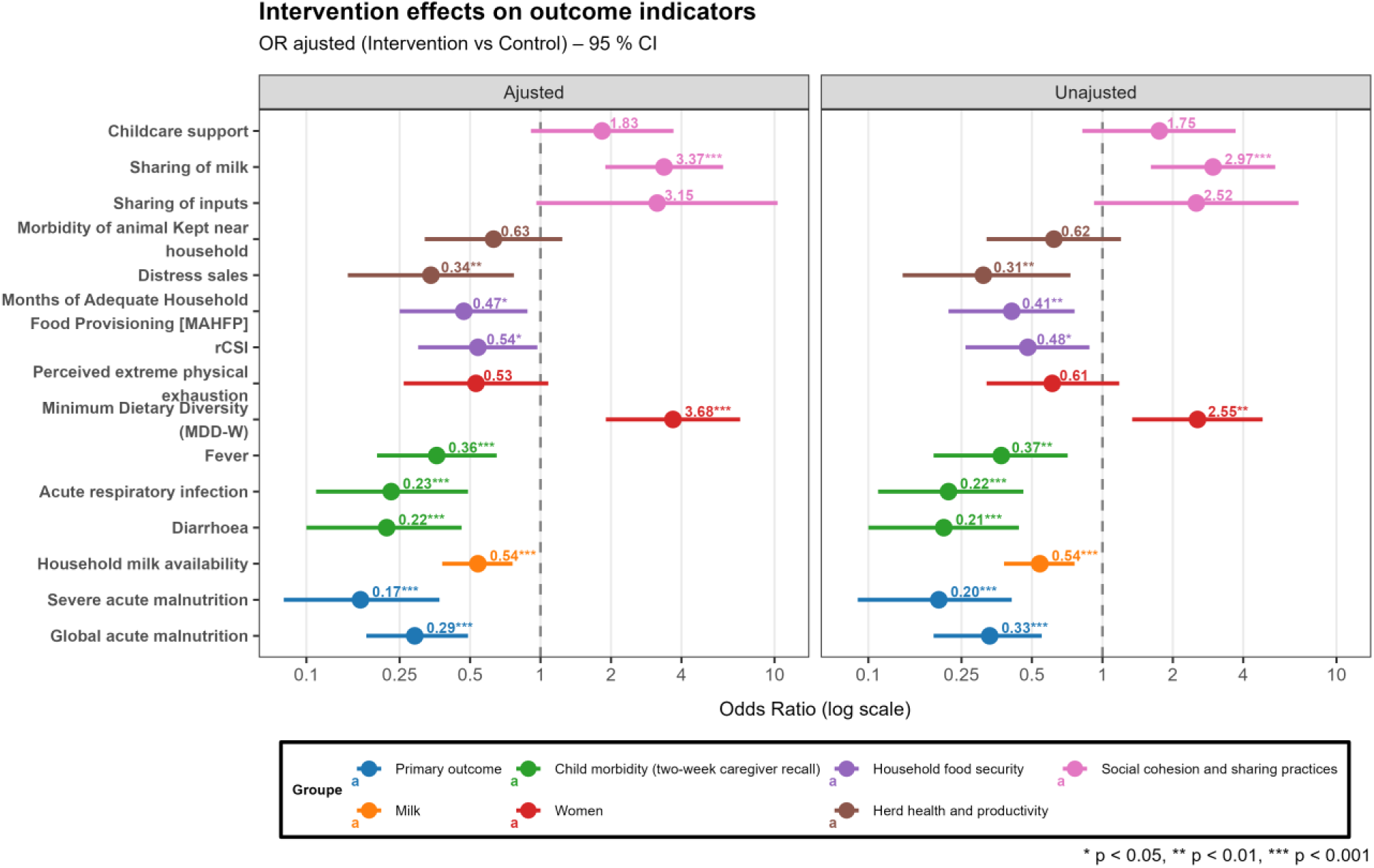
Forest Plot for Primary and Secondary outcomes.

**Table 5:**
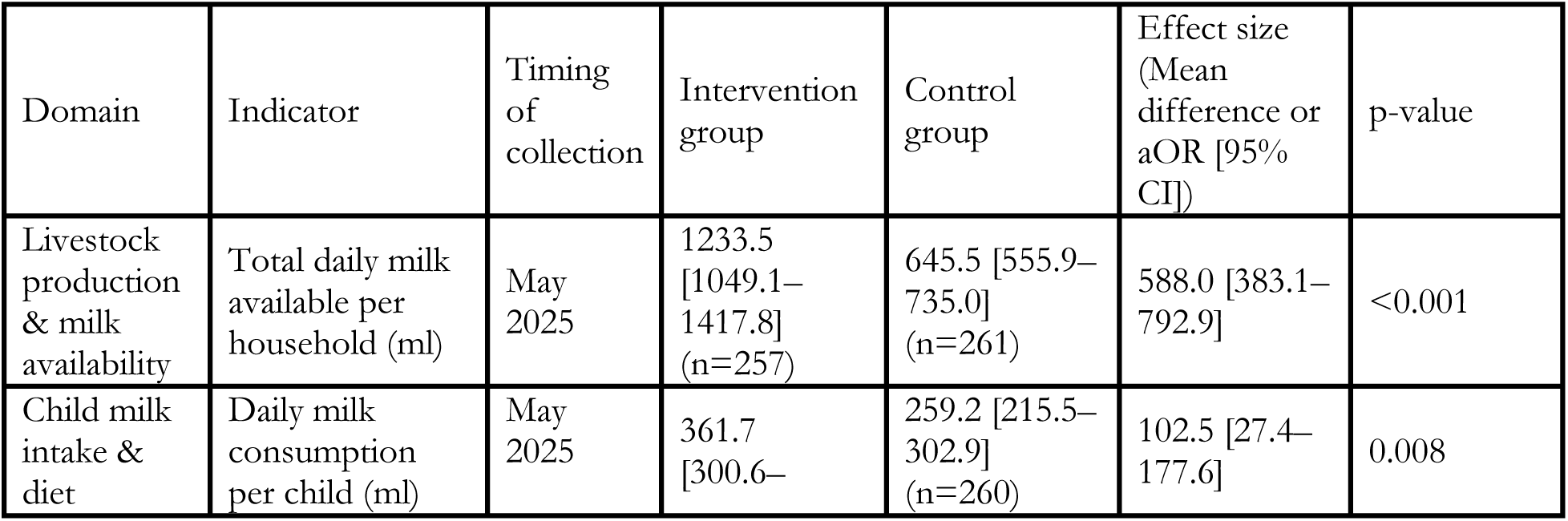
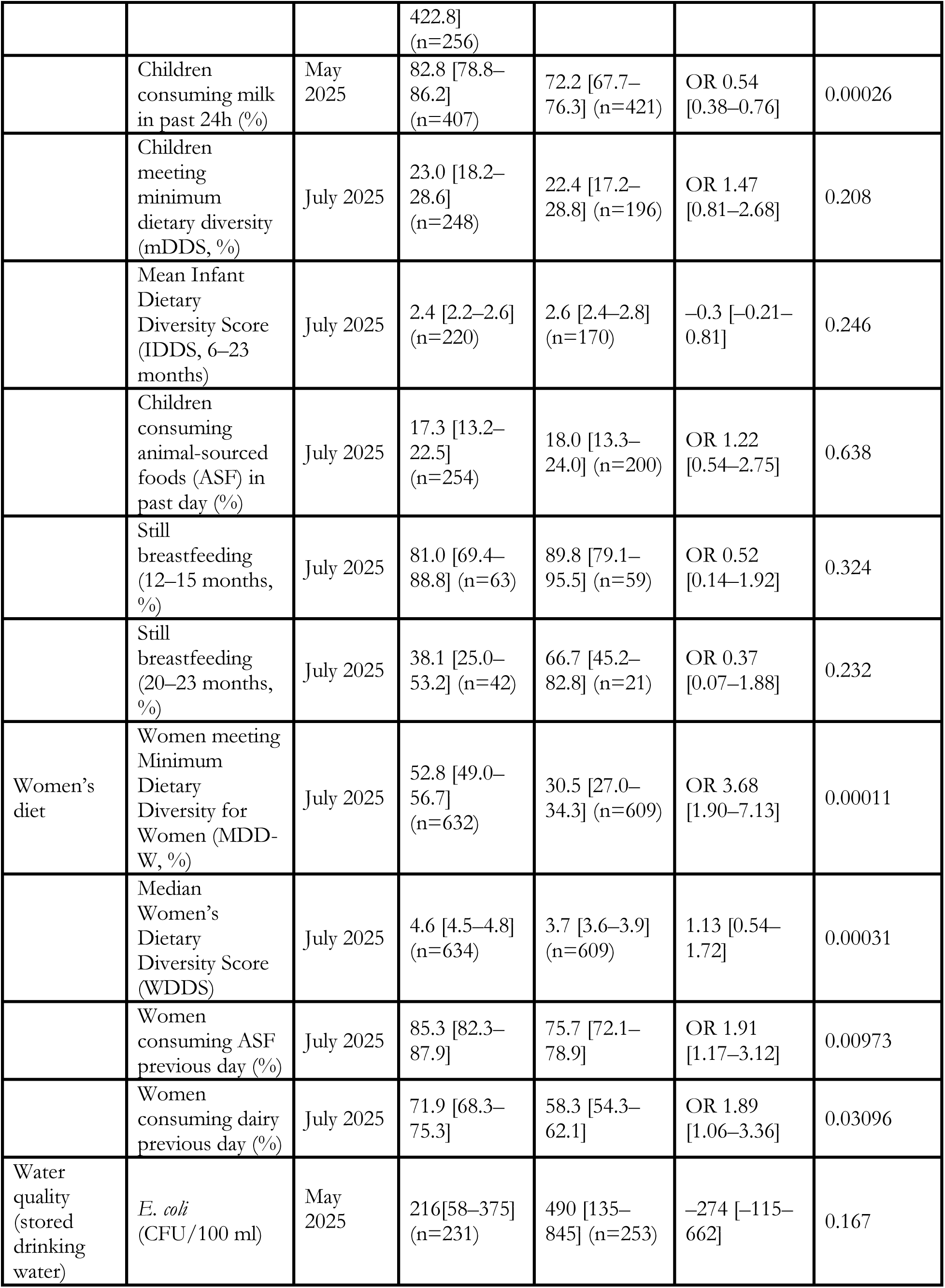
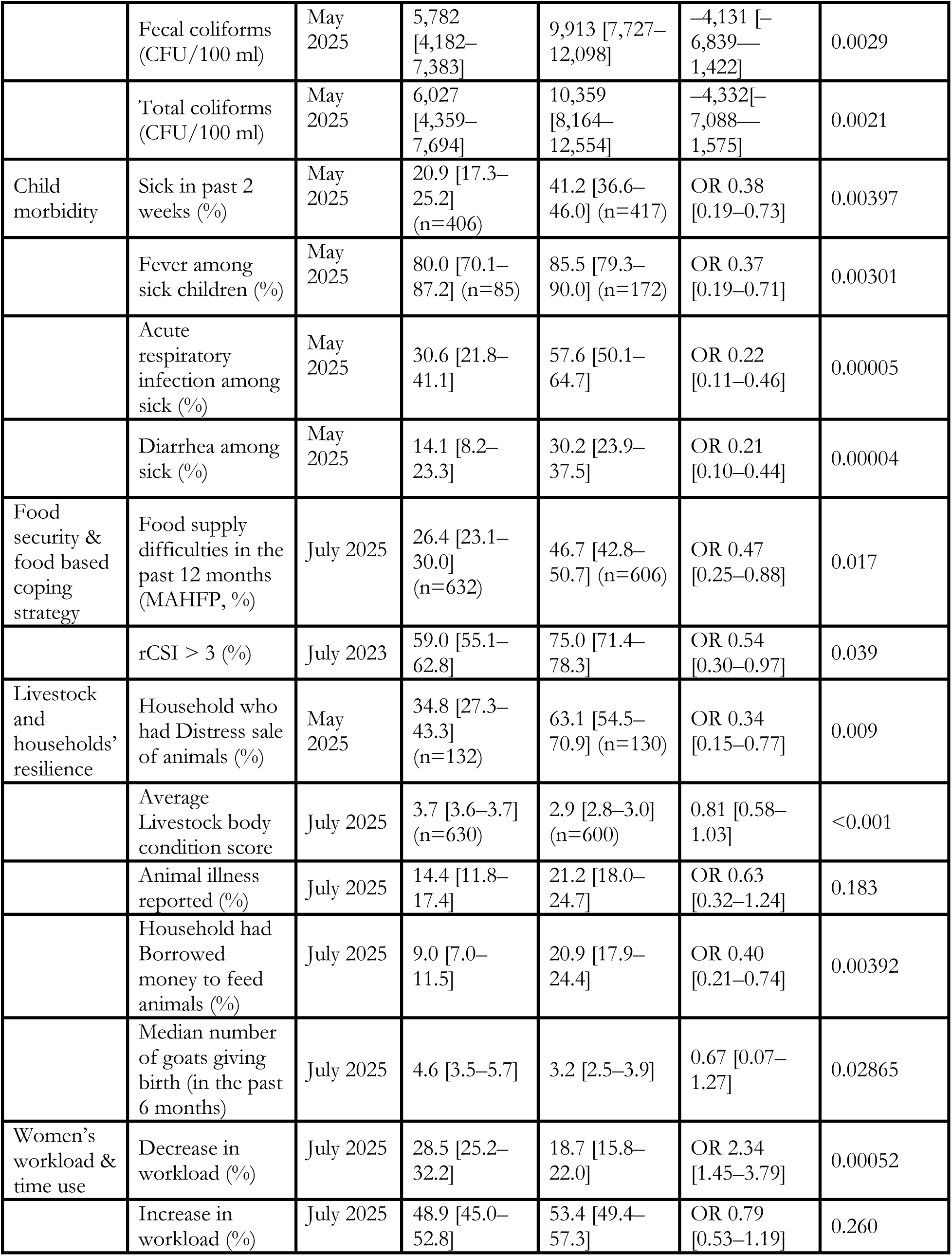
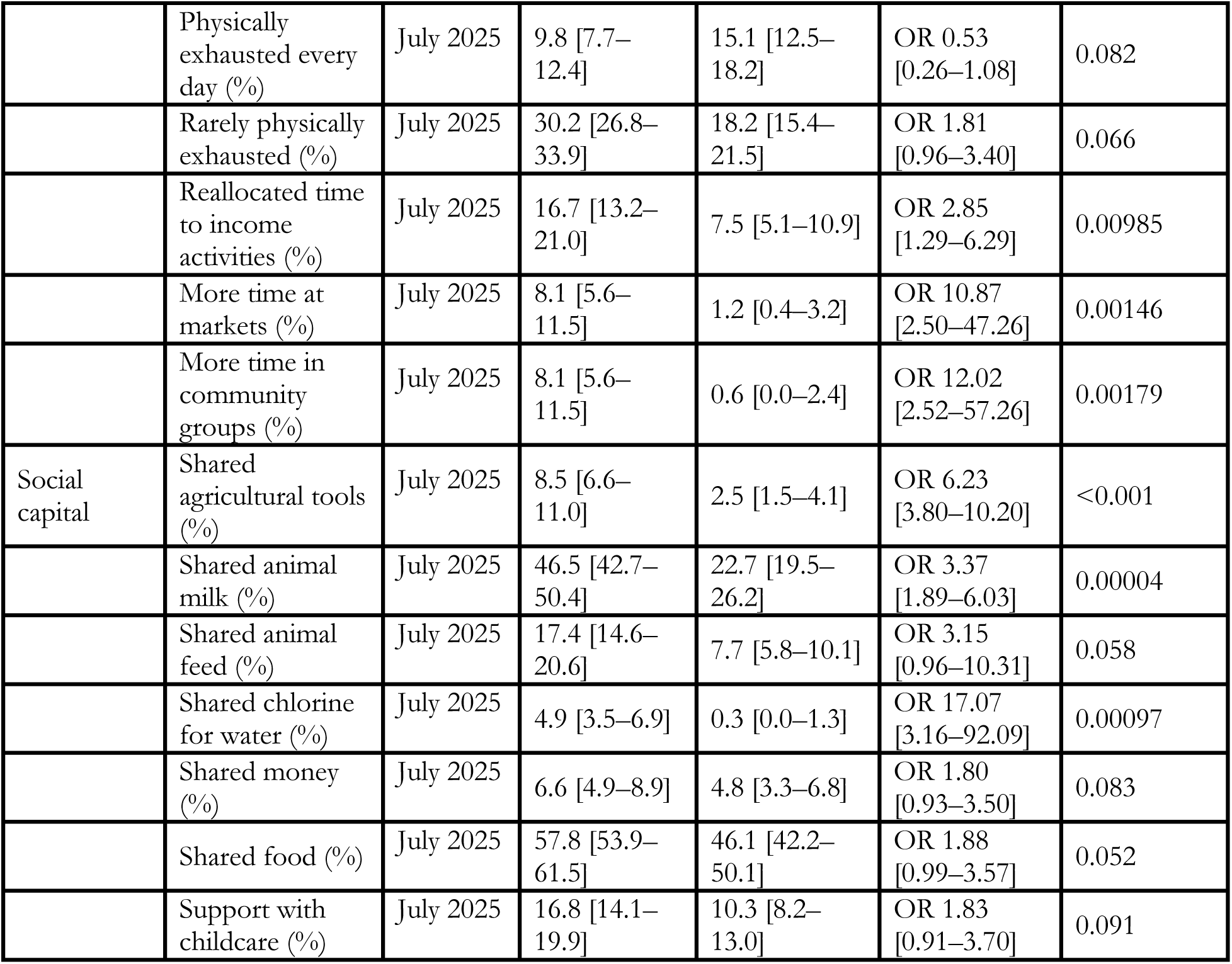
Secondary outcomes.

#### Milk availability and child milk consumption

Intervention households had higher daily milk availability than control households, with a mean difference of +588 mL per household; (p<0·001). Children in intervention households consumed more milk per day on average than the control households (+103 mL per child; p=0·008), and a higher proportion of children consumed milk in the preceding 24h in May 2025 (p=0·00026).

#### Child Diet

No statistically significant differences were observed in child dietary diversity, consumption of non-dairy animal-source foods, or breastfeeding practices measured over the past 24 hours assessed end of July 2025 (all p ≥ 0·208).

#### Child morbidity

Reported child morbidity during the two weeks preceding assessment was lower in the intervention villages. The number of children reported to have suffered from diarrhea, fever or acute respiratory infection over the past 14 days was significantly lower in the intervention villages The adjusted odds of diarrhea or acute respiratory infection were substantially lower in intervention villages than in control villages ( p=0·00005 for acute respiratory infections; p=0·00004 for diarrhea). Fever was significantly less frequent among sick children in intervention villages (p=0·00301)

#### Water quality

Microbiological testing of household drinking water showed lower concentrations of faecal coliforms (p = 0·0029) and total coliforms (p = 0·0021) in intervention households compared with control households at May follow-up. Differences in *E. coli* concentrations did not differ statistically between groups (p = 0·167).

#### Women’s diet and workload

Women in intervention households had higher odds ( p<0·001) of meeting minimum dietary diversity compared with women in control households. Intervention households also reported higher consumption of animal source foods (p=0·00973) and dairy products among women (p=0·03096).

Women in intervention households more frequently reported reductions in workload (p<0·001) and greater engagement in income-generating (p = 0·00985), market (p = 0·00146), and community activities (p = 0·00179).

#### Household food security and livestock

Household food insecurity indicators improved in intervention villages in comparison with control group, with lower odds of food supply difficulties and reduced reliance on negative food-based coping strategies measured by the MAHFP (p=0·017) and rCSI<3 (p=0.039). Intervention households reported fewer distress livestock sales (p=0·009) and higher livestock body condition scores (mean difference +0·81; p<0·001) at follow-up. Reproductive performance also improved, with a higher median number of goats giving birth in the preceding six months (p=0·02865). seasonal timing and short follow-up precluded assessment of reproductive outcomes in other species.

#### Social capital and sharing practices

Measures of social cohesion differed between groups at July follow-up. Households in intervention villages more frequently reported sharing agricultural tools (p < 0·001), animal milk (p = 0·00004), and water treatment supplies (p = 0·00097). Sharing of animal feed (p = 0·058), food (p = 0·052), money (p = 0·083), and childcare support (p = 0·091) was higher in intervention villages but did not differ statistically between groups.

### Return on Investment

A societal benefit cost analysis was conducted over the six-month intervention period (December 2024–May 2025), with projections to 12, 18, and 24 months. Economic returns over the six-month horizon were estimated using observed trial outcomes, while longer-term returns were projected using conservative model-based assumptions.

For the six-month period, total programme costs for 520 households were estimated at USD 193,816 under the base-case scenario. Total monetized benefits over the same period were estimated at USD 1,047,328, corresponding to a benefit cost ratio of 5.4. Under alternative assumptions,^8^ the benefit–cost ratio ranged from 4.7 in the worst-case scenario to 6.5 in the best-case scenario.

When projected beyond the immediate intervention period, estimated returns increased over time. The benefit–cost ratio reached 8.9 at 12 months, 12.4 at 18 months, and 16.4 at 24 months. Increases in projected benefits were primarily driven by reductions in child morbidity and mortality, avoided health-care costs, and improvements in herds productivity and household food security.

Detailed parameter values and sensitivity analyses are presented in the appendix.

**Table 6:** Parameters used for benefit valorization (USD 2025)

| Intervention cost and benefit | 6-months horizon |  |  | 12-months horizon | 18-months horizon | 24-months horizon |
| --- | --- | --- | --- | --- | --- | --- |
| In USD | Base case | Best case | Worst case | Base case | Base case | Base case |

<sup>8</sup> For parameters related to disease prevalence, the lower and upper bounds of the confidence intervals were used to represent the best- and worst-case scenarios, respectively. Most financial parameters were varied by –25 percent (best case) and +25 percent (worst case) relative to their baseline values. For other parameters, we applied the first (Q1) and third (Q3) quartiles of their interquartile confidence intervals to define the range of variation.

|  |  |  |  |  |  |  |
| --- | --- | --- | --- | --- | --- | --- |
| Total costs related to all intervention for 520 households | 193 816 | 145 783 | 243 837 | 193 816 | 193 816 | 193 816 |
| 1. Averted healthcare costs from improved nutritional status in children under five | 57 734 | 41 594 | 75 013 | 115 469 | 124 781 | 230 937 |
| 2. Avoided premature mortality due to reduced acute malnutrition in children under five | 462 485 | 439 711 | 485 260 | 924 971 | 1 319 132 | 1 849 941 |
| 3. Averted healthcare costs from reduced diarrhoea in children under five | 13 041 | 10 184 | 15 629 | 26 082 | 20 368 | 52 163 |
| 4. Averted healthcare costs from reduced acute respiratory infections in children under five | 26 406 | 16 544 | 38 988 | 52 812 | 49 631 | 105 625 |
| 5. Averted veterinary costs from improved livestock morbidity | 791 | 593 | 989 | 791 | 593 | 791 |
| 6. Reduced livestock feed expenses | 62 396 | 46 797 | 77 995 | 62 396 | 46 797 | 62 396 |
| 7. Increased household livestock capital | 376 869 | 352 126 | 403 935 | 449 785 | 497 764 | 692 947 |
| 8. Improved household food security | 47 605 | 35 704 | 59 506 | 95 210 | 107 111 | 190 419 |
| Total benefits for 520 household | 1 047 328 | 943 253 | 1 157 315 | 1 727 515 | 2 166 178 | 3 185 220 |
| Cost–Benefit Ratio (BCR) | 5.4 | 6.5 | 4.7 | 8.9 | 12.4 | 16.4 |

## DISCUSSION

In this cluster-randomised trial conducted in pastoral and agro-pastoral communities in Chad, an integrated livestock management intervention implemented during the pastoral lean season was associated with significantly large reductions in global and severe acute malnutrition at the dry-season peak. The magnitude of the observed effects at the period of highest risk, alongside improvements in intermediate nutrition, health, livelihood, and economic outcomes, provides experimental evidence that anticipatory, livelihood-grounded interventions can prevent predictable seasonal peaks of acute malnutrition in dryland humanitarian settings.

These findings address a central evidence gap highlighted in the WHO guideline on the prevention and management of wasting: the limited availability of rigorously evaluated preventive interventions that reduce acute malnutrition itself, rather than improving only intermediate determinants [32, 33]. Most nutrition-sensitive approaches have demonstrated modest or inconsistent effects on acute malnutrition [34, 35]. By contrast, this trial demonstrates that acting upstream on predictable seasonal risk through an integrated package can substantially reduce both GAM and SAM during a defined high-risk window.

The intervention was explicitly designed to operate through pathways embedded in dryland livelihood systems. Sustaining livestock productivity and household milk availability during the dry season, when milk is scarce, is plausibly linked to improved child dietary intake [36–38]. Concurrent reductions in reported child morbidity and improvements in household drinking water quality are consistent with reduced infection-related nutritional losses, which frequently undermine nutritional gains of food-based interventions in pastoral settings [39–42]. Improvements in women’s dietary diversity and reported workload further suggest that the intervention may have alleviated constraints on caregiving and feeding practices [43, 44]. Although the relative contribution of individual pathways cannot be disentangled, the convergence of observed outcomes is consistent with synergistic and potentially multiplicative effects operating through interconnected mechanisms embedded within local livelihood systems. Importantly, the intervention advances multisectoral programming beyond conventional sector-based integration. Rather than simply co-locating or layering nutrition, WASH, and livestock activities, the intervention design was informed by qualitative research that explored the relational dynamics of livelihood systems and the lived realities of pastoral and agro-pastoral communities. This perspective acknowledges that risks to child nutrition arise through interconnected livelihood practices, where animal health, water access, food production, and caregiving environments jointly shape nutritional outcomes. By anchoring the intervention in seasonal livestock management, caregiving constrains, and human–animal–environment interactions, this approach foregrounds the relational dynamics that shape household vulnerability. In doing so, it operationalises multisectorality at the level where households and communities experience risk through interconnected livelihood, care and ecological systems, rather than through fragmented, sector-specific delivery mechanisms.

The large effect sizes observed in this trial should be interpreted in light of the known seasonal patterns of acute malnutrition. Prevalence of acute malnutrition at the dry-season peak in drylands is consistently extreme, reflecting a concentrated period of vulnerability linked to environmental stress, livelihood dynamics, and infection risk [45]. Intervening during this narrow but predictable window may amplify observable impacts relative to interventions implemented outside peak-risk periods. The dry-season peak observed in Chad mirrors patterns documented across many drylands’ contexts [46]. However, effect sizes may differ in lower-risk contexts or in settings with less pronounced seasonality, underscoring the importance of contextual adaptation.

The intervention generated substantial economic returns, with benefit–cost ratios estimated over the six-month trial horizon and projected to increase over longer time frames. These returns were primarily driven by avoided healthcare costs, reductions in child morbidity and mortality, and sustained livestock productivity.

The findings underscore the potential of preventive, livelihood-based approaches as cost-efficient complements to reactive nutrition interventions. Crucially, these results indicate that integrated anticipatory interventions not only save lives and reduce future caseloads of acute malnutrition, but also improve the efficiency of scarce humanitarian resources.

Several limitations warrant consideration. First, the integrated nature of the intervention precludes attribution of effects to individual components. Second, follow-up beyond the immediate post-peak period was limited, restricting inference on longer-term nutritional trajectories. Third, the trial was conducted in a specific dryland context characterized by pastoral livelihoods and pronounced seasonality, which may limit generalizability to other settings. Finally, although efforts were made to minimise measurement bias, blinding of participants and implementers was not feasible.

## CONCLUSION

In pastoral and agro-pastoral drylands, child acute malnutrition follows predictable seasonal patterns shaped by livelihood dynamics, infection risks at the human–animal–environment interface, and caregiving constraints. This cluster-randomized controlled trial provides experimental evidence that an anticipatory, integrated livestock management intervention implemented ahead of the pastoral lean season can prevent seasonal peaks of acute malnutrition and reduce child morbidity, while mitigating women’s workload, reinforcing household resilience and generating substantial economic returns.

This study shows that context-specific livelihood-based interventions, when integrated with One Health–informed risk mitigation, can reduce the prevalence of acute malnutrition and lessen the need for purely reactive interventions after deterioration has occurs.

In dryland settings characterized by recurrent seasonal peaks of acute malnutrition and in a context of constrained humanitarian financing, anticipatory integrated approaches that act on underlying livelihood- and infection-related risk pathways could represent an effective strategy to prevent child acute malnutrition.

## Data Availability

All data produced in the present study are available upon reasonable request to the authors

## Trial registration

Pan African Clinical Trials Registry (PACTR202504882709268).

The authors have declared no competing interest.

## APPENDIX

## Appendix 1

### Methods for Return of Investment study

The economic evaluation aimed to assess the return on investment (ROI) of the integrated livestock management in comparison to a control group. A benefit–cost analysis was conducted to estimate the intervention gains relative to its total implementation costs. The analysis used a representative subsample of 520 households drawn equally from 26 control villages and 26 intervention villages.

#### Time horizon

The primary analysis covered a six-month period (December 2024 – May 2025), corresponding to the project’s implementation phase. To capture sustained impacts on selected secondary outcomes, additional analyses were conducted using extended time horizons of 12, 18, and 24 months.

#### Analysis perspective

The analysis adopted a societal perspective, encompassing all costs and benefits associated with the interventions; whether borne by project implementers or by participating households. This approach ensured that both financial expenditures and household opportunity costs were reflected in the overall cost-benefit balance.

#### Intervention cost: data collection and analysis

An activity-based costing approach was applied to estimate the intervention costs. All activities were disaggregated into their component inputs and costs were estimated for each input. Programme staff were consulted to develop a comprehensive and mutually exclusive list of cost centres corresponding to each intervention component (Table 7), ensuring complete coverage of implementation activities and avoiding double-counting.

**Table 7:** Cost centres and their description.

| Cost centres | Description |
| --- | --- |
| Training related to input distribution and SBC counselling activities | Includes expenses related to staff salaries, per diems, transport, and supplies for training activities on SBC sensitization and input distribution. |
| Distributed inputs | This includes the costs associated with the distributed inputs (cottonseed cake, maize bran, hay, and salt licks, soap and Chlorine), including the expenses related to their purchase, quality control, storage, and transportation. It also accounts for personnel time costs incurred during procurement and distribution activities. |
| Households' costs | This includes the time spent by the households attending sensitization activities and distribution of inputs |

It was assumed that no cost differences existed between the intervention and comparison groups for activities common to both groups such as routine vaccination and deworming. Accordingly, the analysis focused on incremental costs directly attributable to the intervention. Routine operational and research related expenditures were excluded from the ROI analysis. For the supplementary analyses extending beyond the implementation period, it was likewise assumed that no additional cost differences would arise between the groups.

All costs were first calculated in local currency (*Communauté Financière Africaine* [CFA] francs) and converted to the current 2025 USD using the official exchange rate of *June* 2025. No discounting was applied, as the main analytical time horizon was less than one year.

All financial data were obtained from the project’s financial records. Information on time allocation was primarily collected through key informant interviews with project staff. To estimate the opportunity cost of time devoted by households to project activities, the human capital approach was applied. The value of participants’ time was monetized using the guaranteed minimum agricultural wage in Chad as defined by national labor regulations [1].

#### Intervention benefits: data collection and analysis

The analysis captured all monetized benefits generated by integrated livestock management interventions. Table 8 summarizes the specific benefit categories, indicators, and valuation methods used. The value of benefits was calculated by aggregating all individual benefits that met two criteria:

- They showed a statistically significant difference between intervention and control groups
- They could be quantified and expressed in monetary terms.

**Table 8:**
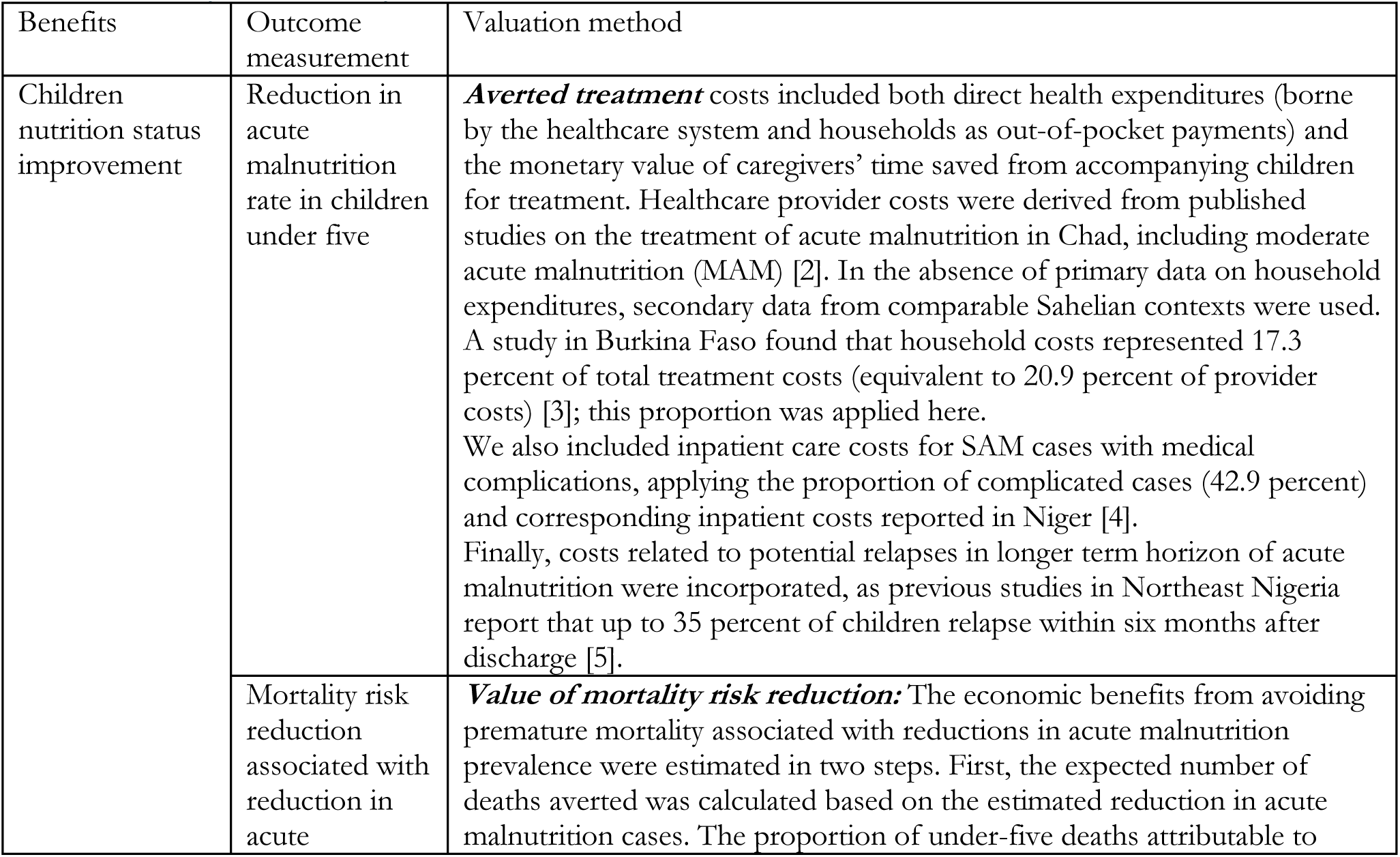
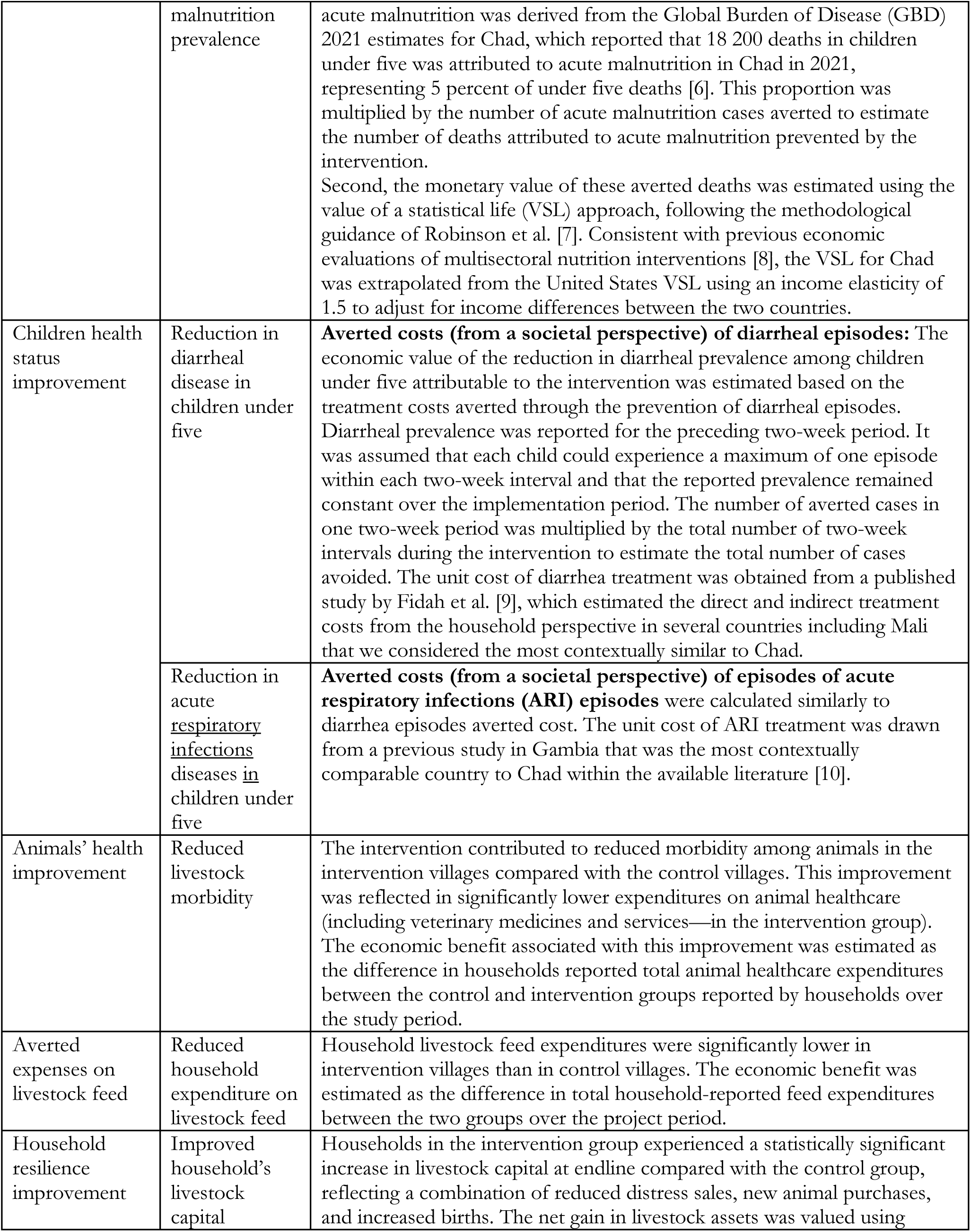
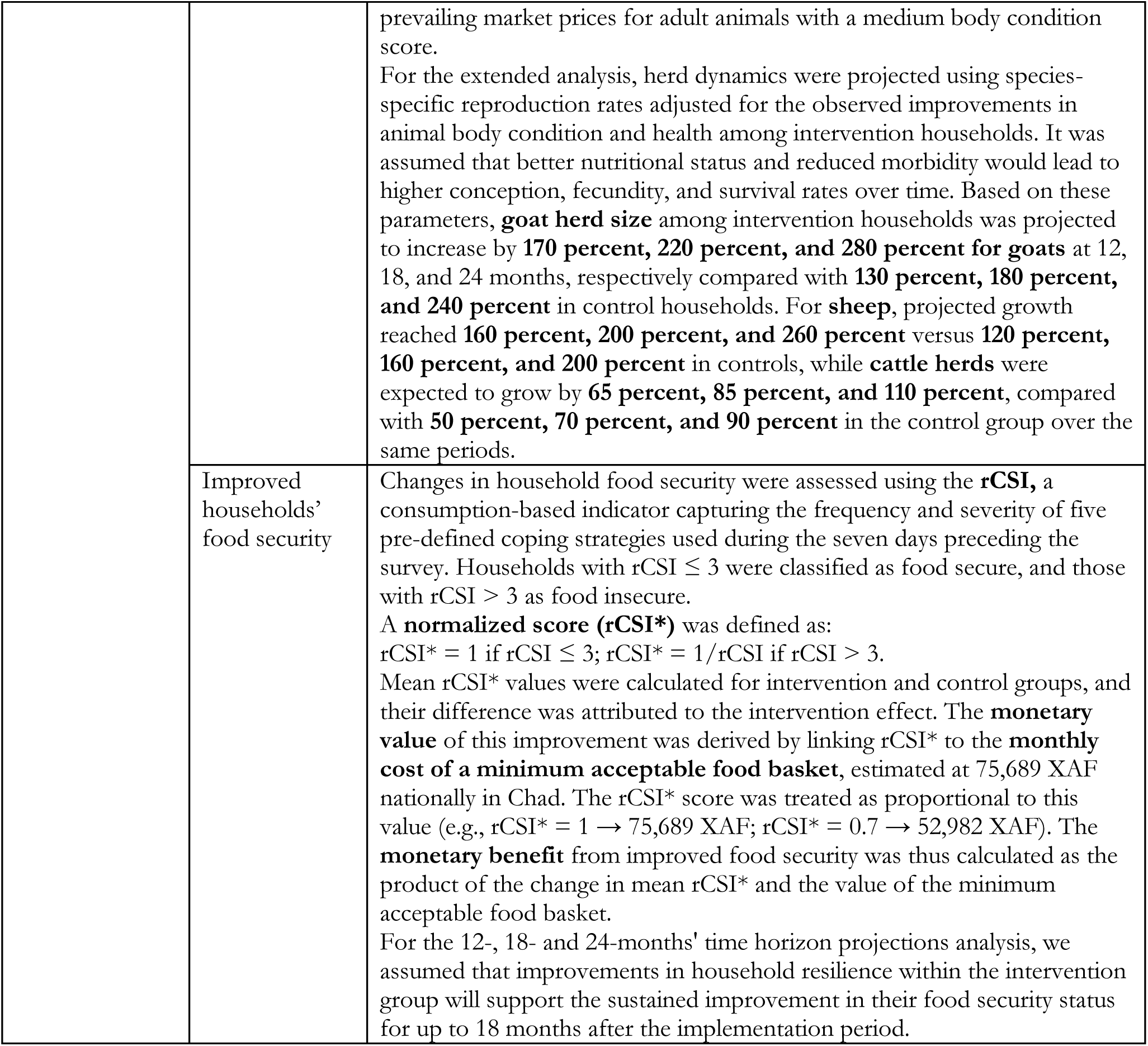
Benefits and methods for their measurement and valuation.

| Benefits | Outcome measurement | Valuation method |
| --- | --- | --- |
| Children nutrition status improvement | Reduction in acute malnutrition rate in children under five | **Averted treatment** costs included both direct health expenditures (borne by the healthcare system and households as out-of-pocket payments) and the monetary value of caregivers' time saved from accompanying children for treatment. Healthcare provider costs were derived from published studies on the treatment of acute malnutrition in Chad, including moderate acute malnutrition (MAM) [2]. In the absence of primary data on household expenditures, secondary data from comparable Sahelian contexts were used. A study in Burkina Faso found that household costs represented 17.3 percent of total treatment costs (equivalent to 20.9 percent of provider costs) [3]; this proportion was applied here. We also included inpatient care costs for SAM cases with medical complications, applying the proportion of complicated cases (42.9 percent) and corresponding inpatient costs reported in Niger [4]. Finally, costs related to potential relapses in longer term horizon of acute malnutrition were incorporated, as previous studies in Northeast Nigeria report that up to 35 percent of children relapse within six months after discharge [5]. |
|  | Mortality risk reduction associated with reduction in acute | **Value of mortality risk reduction:** The economic benefits from avoiding premature mortality associated with reductions in acute malnutrition prevalence were estimated in two steps. First, the expected number of deaths averted was calculated based on the estimated reduction in acute malnutrition cases. The proportion of under-five deaths attributable to |
|  | malnutrition prevalence | acute malnutrition was derived from the Global Burden of Disease (GBD) 2021 estimates for Chad, which reported that 18 200 deaths in children under five was attributed to acute malnutrition in Chad in 2021, representing 5 percent of under five deaths [6]. This proportion was multiplied by the number of acute malnutrition cases averted to estimate the number of deaths attributed to acute malnutrition prevented by the intervention.<br>Second, the monetary value of these averted deaths was estimated using the value of a statistical life (VSL) approach, following the methodological guidance of Robinson et al. [7]. Consistent with previous economic evaluations of multisectoral nutrition interventions [8], the VSL for Chad was extrapolated from the United States VSL using an income elasticity of 1.5 to adjust for income differences between the two countries. |
| Children health status improvement | Reduction in diarrheal disease in children under five | **Averted costs (from a societal perspective) of diarrheal episodes:** The economic value of the reduction in diarrheal prevalence among children under five attributable to the intervention was estimated based on the treatment costs averted through the prevention of diarrheal episodes. Diarrheal prevalence was reported for the preceding two-week period. It was assumed that each child could experience a maximum of one episode within each two-week interval and that the reported prevalence remained constant over the implementation period. The number of averted cases in one two-week period was multiplied by the total number of two-week intervals during the intervention to estimate the total number of cases avoided. The unit cost of diarrhea treatment was obtained from a published study by Fidah et al. [9], which estimated the direct and indirect treatment costs from the household perspective in several countries including Mali that we considered the most contextually similar to Chad. |
|  | Reduction in acute <u>respiratory infections</u> diseases in children under five | **Averted costs (from a societal perspective) of episodes of acute respiratory infections (ARI) episodes** were calculated similarly to diarrhea episodes averted cost. The unit cost of ARI treatment was drawn from a previous study in Gambia that was the most contextually comparable country to Chad within the available literature [10]. |
| Animals' health improvement | Reduced livestock morbidity | The intervention contributed to reduced morbidity among animals in the intervention villages compared with the control villages. This improvement was reflected in significantly lower expenditures on animal healthcare (including veterinary medicines and services—in the intervention group). The economic benefit associated with this improvement was estimated as the difference in households reported total animal healthcare expenditures between the control and intervention groups reported by households over the study period. |
| Averted expenses on livestock feed | Reduced household expenditure on livestock feed | Household livestock feed expenditures were significantly lower in intervention villages than in control villages. The economic benefit was estimated as the difference in total household-reported feed expenditures between the two groups over the project period. |
| Household resilience improvement | Improved household's livestock capital | Households in the intervention group experienced a statistically significant increase in livestock capital at endline compared with the control group, reflecting a combination of reduced distress sales, new animal purchases, and increased births. The net gain in livestock assets was valued using |
|  |  | prevailing market prices for adult animals with a medium body condition score.<br>For the extended analysis, herd dynamics were projected using species-specific reproduction rates adjusted for the observed improvements in animal body condition and health among intervention households. It was assumed that better nutritional status and reduced morbidity would lead to higher conception, fecundity, and survival rates over time. Based on these parameters, **goat herd size** among intervention households was projected to increase by **170 percent, 220 percent, and 280 percent for goats** at 12, 18, and 24 months, respectively compared with **130 percent, 180 percent, and 240 percent** in control households. For **sheep**, projected growth reached **160 percent, 200 percent, and 260 percent** versus **120 percent, 160 percent, and 200 percent** in controls, while **cattle herds** were expected to grow by **65 percent, 85 percent, and 110 percent**, compared with **50 percent, 70 percent, and 90 percent** in the control group over the same periods. |
|  | Improved households' food security | Changes in household food security were assessed using the **rCSI**, a consumption-based indicator capturing the frequency and severity of five pre-defined coping strategies used during the seven days preceding the survey. Households with $rCSI \leq 3$ were classified as food secure, and those with $rCSI > 3$ as food insecure.<br>A **normalized score (rCSI\*)** was defined as:<br>$rCSI^* = 1$ if $rCSI \leq 3$; $rCSI^* = 1/rCSI$ if $rCSI > 3$.<br>Mean $rCSI^*$ values were calculated for intervention and control groups, and their difference was attributed to the intervention effect. The **monetary value** of this improvement was derived by linking $rCSI^*$ to the **monthly cost of a minimum acceptable food basket**, estimated at 75,689 XAF nationally in Chad. The $rCSI^*$ score was treated as proportional to this value (e.g., $rCSI^* = 1 \rightarrow 75,689$ XAF; $rCSI^* = 0.7 \rightarrow 52,982$ XAF). The **monetary benefit** from improved food security was thus calculated as the product of the change in mean $rCSI^*$ and the value of the minimum acceptable food basket.<br>For the 12-, 18- and 24-months' time horizon projections analysis, we assumed that improvements in household resilience within the intervention group will support the sustained improvement in their food security status for up to 18 months after the implementation period. |

As with costs, all benefits were valued at 2025 USD.

#### Return on investment outcome

The return on investment was evaluated using the benefit–cost ratio (BCR). The BCR represents the ratio of the present value of total benefits to the present value of total costs. A BCR greater than one suggests that the intervention generated positive net economic returns over the analysis period. The BCR also serves as an indicator of efficiency, quantifying the monetary value of benefits generated per USD invested.

#### Sensitivity analysis

We conducted a multi-way deterministic sensitivity analysis to assess how variations in input parameters affect the BCR. In this analysis, multiple parameters were varied simultaneously within predefined plausible ranges. For parameters related to disease prevalence, the lower and upper bounds of the confidence intervals were used to represent the best- and worst-case scenarios, respectively. Most financial parameters were varied by −25 percent (best case) and +25 percent (worst case) relative to their baseline values. For other parameters, we applied the first (Q1) and third (Q3) quartiles of their interquartile confidence intervals to define the range of variation.

#### Project cost

The total implementation cost of the intervention was calculated based on all activities carried out during the project period. Table 9 provides a detailed breakdown of these costs by activity, along with the corresponding cost per household from the provider perspective. The overall **cost per household** was **USD 370 from**

**Table 9:** Breakdown of project implementation costs by activity and cost per household from provider perspectives.

| Cost item | cost (USD) | Cost per household (USD) |
| --- | --- | --- |
| Purchase and quality control of cottonseed cake (per kg) | 0.49 | 62 |
| Purchase and quality control of maize bran (per kg) | 0.46 | 136 |
| Purchase and quality control of mineral lick block (5 kg) | 3.02 | 3 |
| Purchase and quality control of hay (5 kg) | 4.85 | 48 |
| Purchase of chlorine bottle (1 l) | 1.83 | 4 |
| Purchase of soap (one bar) | 0.35 | 3 |
| Training of ASRADD for distribution and awareness-raising | 2 870.67 | 3 |
| Vehicle rental for six awareness sessions for 755 households in 38 villages | 10 275.60 | 12 |
| Vehicle rental for ten distribution rounds & awareness recall for 755 households in 38 villages | 14 271.67 | 17 |
| Fuel for six awareness sessions for 755 households in 38 villages | 17 779.66 | 22 |
| Fuel for ten rounds of feed & soap distribution and awareness recall for 755 households in 38 villages | 8 466.50 | 11 |
| Human resources ASRADD – chlorine distribution and six awareness sessions for 755 households in 38 villages | 11 417.34 | 14 |
| Human resources ASRADD – for ten rounds of feed and soap distribution and recall of awareness session for 755 households in 38 villages | 20 301.45 | 24 |
| Targeting beneficiaries' households within 38 villages | 10 309.50 | 11 |
| Total cost by household in USD |  | 370 |

#### provider perspective

This estimate excludes the cost of the initial formative study (to be conducted in each new context) and FAO’s overall supervision and evaluation costs.

From household perspective, the cost borne by beneficiaries was estimated at USD 3.25 per household, reflecting the time spent attending counselling sessions, managing livestock (feeding and milking), and receiving inputs such as soap and water-treatment products. Counselling sessions were deliberately held early in the morning or in the evening to minimise disruption to households’ usual economic and livelihood activities. Qualitative respondents reported that receiving feed directly at home saved them the time they would normally spend purchasing feed in markets or collecting fodder. Beneficiaries also noted that the intervention did not increase the time required for feeding or milking their animals.

For the **520 households** included in the ROI study, total costs were assessed under three analytical perspectives: the provider perspectives (implementation and delivery), the household perspective (time and resources contribution by beneficiaries), and the societal perspective (the combined value of all costs incurred by both implementers and participating households) (Table 10).

**Table 10:** Total project costs by analytical perspective and scenario (USD 2025)

| Perspective |  | Base case | Best case | Worst case |
| --- | --- | --- | --- | --- |
| Provider perspective |  | 192 131 | 144 098 | 241 203 |
| Household perspective |  | 1 685 | 1 685 | 2 633 |
| Societal perspective | (combined) | 193 816 | 145 783 | 243 837 |

#### Benefit valorization results

Table 11 presents the parameters and unit values used to monetize the benefits generated by the intervention under base, best, and worst-case scenarios.

**Table 11:** Parameters used for benefit valorization at six months (USD 2025)

| Benefit component | Parameter | Base case | Best case | Worst case |
| --- | --- | --- | --- | --- |
| 1. Averted healthcare costs – improved nutritional status in children under five | Average cost of treating an acute malnutrition episode (including MAM management), provider perspective | 130.65 | 97.99 | 163.31 |
|  | Cost of treatment, household perspective (out-of-pocket and caregivers' opportunity cost) | 27.31 | 20.48 | 34.13 |
| 2. Avoided premature mortality – reduced acute malnutrition in children under five | Estimated number of child deaths averted (520 households) | 14 | 13 | 15 |
| | Value of a statistical life (VSL) in Chad, 2015 international \$ | 75 395 | 75 395 | 75 395 |
| 3. Averted healthcare costs – reduced diarrhea in children under five | Average treatment cost per episode, household perspective | 22.50 | 16.88 | 28.13 |
| 4. Averted healthcare costs – reduced acute respiratory infections in children under five | Average treatment cost per episode, household perspective | 6.6 | 3.2 | 12.4 |
| 5. Averted veterinary costs – improved livestock morbidity | Reduction in household expenditure on veterinary medicines/services vs control | 1.7 | 1.3 | 2.1 |
| 6. Reduced livestock feed expenses | Feed cost savings per household (dry season, over six months) | 119.99 | 89.99 | 149.99 |
| 7. Increased household livestock capital | Market value of a goat | 38 | 34 | 42 |
|  | Market value of a sheep | 84 | 79 | 91 |
|  | Market value of a cow | 324 | 304 | 344 |
|  | Market value of net livestock capital gain among intervention households (over six months) | 725 | 677 | 777 |
| 8. Improved household food security | Variation in normalized rCSI score attributable to the intervention | 0.113 | 0.113 | 0.113 |
|  | Monthly cost of a minimum acceptable food basket | 135 | 101 | 169 |
|  | Monetary value of food-security gains for intervention group over six months | 98.7 | 74 | 123 |

## Footnotes

1 The humanitarian reset refers to a proposed overhaul of the humanitarian aid system aimed at improving efficiency and effectiveness in response to “a profound crisis of legitimacy, morale, and funding”. The reset was instigated by Tom Fletcher, the Emergency Relief Chief on 10 March 2025. Source available at The humanitarian reset (10 March 2025) | OCHA

2 Seasonal access to milk is strongly influenced by herd composition, as well as the availability of complementary feed when pasture resources become scarce. Local markets offer limited options for animal feed, and while communities do purchase feed during dry season, high market prices constrain both the quantity and quality they can access.

3 A recent FAO-led study in the study area Kanem and Bar El Gazel, Chad found that 36.8% of children under five were infected with *Campylobacter*, 12.3% with *Cryptosporidium parvum*, and that 43.3% of household water sources were contaminated with *Salmonella*. Reference: From Herds to Child health: An integrated livestock management intervention to prevent acute malnutrition in Chad, FAO, Forthcoming 2026

5 Because logistical uncertainty made it unlikely that a full endline could be implemented during the peak malnutrition season, we prioritized capturing peak outcomes with a reduced interim sample and continued the intervention and follow-up thereafter to avoid missing peak effects altogether while still documenting post-peak trajectories

6 A TLU was defined as one cow or camel, or ten sheep and goats

7 It is therefore important to interpret the timing of the intervention within the context of the 2024-2025 relatively favourable climatic conditions, as outcomes related to vegetation and livestock productivity may differ under less favourable rainfall patterns. In the event of an unfavourable preceding rainy season, earlier implementation of the intervention is recommended.

8 For parameters related to disease prevalence, the lower and upper bounds of the confidence intervals were used to represent the best- and worst-case scenarios, respectively. Most financial parameters were varied by −25 percent (best case) and +25 percent (worst case) relative to their baseline values. For other parameters, we applied the first (Q1) and third (Q3) quartiles of their interquartile confidence intervals to define the range of variation.

## REFERENCES

1. United Nations Children’s Fund (UNICEF), World Health Organization, International Bank for Reconstruction and Development/The World Bank. Levels and trends in child malnutrition: UNICEF / WHO / World Bank Group Joint Child Malnutrition Estimates. Key findings of the 2025 edition. Geneva: World Health Organization; 2025. Licence: CC BY-NC-SA 3.0 IGO

2. Young H, Marshak A. Persistent global acute malnutrition: A discussion paper on the scope of the problem, its drivers and strategies for moving forward for policy, practice and research; 2018. Boston, USA, Feinstein International Center, Tufts University

3. Global Nutrition Report. Chad nutrition profile. In: Global Nutrition Report. [Cited 5 November 2025]. https://globalnutritionreport.org/resources/nutrition-profiles/africa/middle-africa/chad/

4. Integrated Food Security Phase Classification (IPC). Chad acute malnutrition analyses and projections, 2020–2025. IPC Global Support Unit; 2021–2025. Available from: https://www.ipcinfo.org/ipc-country-analysis/details-map/en/?iso3=TCD (accessed Nov 5, 2025).

5. Wambua J, Ali A, Ukwizabigira J.B, et al. Prevalence and risk factors of under-five mortality due to severe acute malnutrition in Africa: a systematic review and meta-analysis. Syst Rev 2025;14, 29. 10.1186/s13643-024-02740-9

6. Global Nutrition Cluster. Nutrition Cluster Power BI dashboard: humanitarian nutrition activity data. Global Nutrition Cluster. Available from: https://app.powerbi.com/view?r=eyJrIjoiOTVmMjQ3OWItZWIxZS00YmIzLThjYjktZjZiMDRhZTBmNjcxIiwidCI6IjVlZjFhZDQ4LWJkZTgtNDY0My1hODlhLWVkMTQyNmI0NGJjMyJ9 (accessed Jan 18, 2026).

7. Food Security Cluster. n.d. “Chad: Food Security Cluster Country Dashboard.” FSCluster. Accessed January 18, 2026. https://fscluster.org/chad.

8. ALNAP. Global Humanitarian Assistance 2025. London: ALNAP/ODI; 2025. Available from: https://www.alnap.org/what-we-do/global-humanitarian-assistance (accessed 18 January 2026).

9. Osendarp S, Ruel M, Udomkesmalee E, et al. The full lethal impact of massive cuts to international food aid. Nature, 2025;640(8057): 35–37. 10.1038/d41586-025-00898-3

10. Food and Agriculture Organization of the United Nations (FAO). Twin peaks: The seasonality of acute malnutrition, conflict and environmental factors in Chad, South Sudan and the Sudan. Briefing note: Mind the gap – Bridging the research, practice and policy divide to enhance livelihood resilience in conflict settings. 2019. 1st ed. Rome, FAO. (FAO job no. CA6984EN). http://www.fao.org/documents/card/en/c/ca6984en

11. Venkat A, Marshak A, Young H, Naumova E.N. Seasonality of acute malnutrition in African drylands: Evidence from 15 years of SMART surveys. Food and Nutrition Bulletin, 2023;44(2_suppl): S94–S108. 10.1177/03795721231178344

12. Marshak A, Young H, Naumova E.N. The complexity of the seasonality of nutritional status: Two annual peaks in child wasting in eastern Chad. Food and Nutrition Bulletin, 2023;44(2_suppl): S109–S118. 10.1177/03795721231181715

13. Famine Early Warning Systems Network (FEWS NET). Chad livelihoods seasonal calendar. Washington, DC, FEWS NET. 2024. https://fews.net/west-africa/chad

14. Young H. Nutrition in Africa’s drylands: A conceptual framework for addressing acute malnutrition. 2020. Feinstein International Center Working Paper. Boston, USA, Feinstein International Center, Friedman School of Nutrition Science and Policy, Tufts University.

15. Young H, Osman A, Radday A, et al. Improving the way we address acute malnutrition in Africa’s drylands. Field Exchange, 2021;65: 14. Emergency Nutrition Network (ENN). https://www.ennonline.net/fex/65/en/improving-way-we-address-acute-malnutrition-africas-drylands

16. Young H, Jenkins N, Osman A.M, et al. Introduction: Addressing the basic drivers of acute malnutrition. Food and Nutrition Bulletin, 2023;44(2_suppl): S5–S8. 10.1177/03795721231186709

17. Njinkeu D, Tchana Tchana F, Lohi J. S, Alli M. O. Chad’s Livestock: Securing Cross-Border Value-Chain Post-COVID-19 (Policy Research Working Paper 10830). 2024. World Bank Group. Retrieved from https://documents1.worldbank.org/curated/en/099747206272432864/pdf/IDU1dfabc2e212a60144e919ac1148f07b91d6d6.pdf

18. Muema J, Mutono N, Kisaka S, et al. The impact of livestock interventions on nutritional outcomes of children younger than five years old and women in Africa: A systematic review and meta-analysis. Frontiers in Nutrition, 2023;10: 1166495. 10.3389/fnut.2023.1166495

19. Global Alliance for Improved Nutrition (GAIN). Investing in nutrition, investing in women: Priority value chains in sub-Saharan Africa. 2025. Geneva, GAIN.

20. De Haan C, Dubern E, Garancher B, Quintero C. Pastoralism development in the Sahel: A road to stability? 2016. Washington, DC, World Bank (International Bank for Reconstruction and Development).

21. Baltenweck I, Achandi E.L, Bullock R.M, et al. Livestock as a pathway to women’s empowerment in low-and middle-income countries: A scoping review. The Journal of Development Studies, 2024;60(6): 813–830. 10.1080/00220388.2024.2319072

22. Hira F.T.Z, Alam M.J, Begum I.A. Women’s empowerment in the livestock sector as a tool to enhance child nutrition: A review. Discover Sustainability, 2025;6: 76. 10.1007/s43621-024-00665-w

23. Sadler K, Mitchard E, Abdi A, et al. Milk matters: The impact of dry-season livestock support on milk supply and child nutrition in Somali Region, Ethiopia. 2012. Boston, USA, Feinstein International Center, Tufts University; Save the Children; USAID.

24. Food and Agriculture Organization of the United Nations (FAO). Livestock for Health in Kenya: Contributing to the prevention of acute malnutrition among children in pastoral households through nutrition-sensitive livestock programming in Marsabit County. 2023. Rome, FAO. https://openknowledge.fao.org/server/api/core/bitstreams/047fcd32-57c6-4960-8733-0034f5313a9e/content

25. Luc G, Keita M, Houssoube F, et al. Community clustering of food insecurity and malnutrition associated with systemic drivers in Chad. Food and Nutrition Bulletin, 2023;44(2_suppl): S69–S82. 10.1177/03795721231189970

26. Headey D, Hirvonen K. Is exposure to poultry harmful to child nutrition? An observational analysis for rural Ethiopia. PLoS ONE, 2016;11(8): e0160590. 10.1371/journal.pone.0160590

27. Mokdad A.H, Khalil I.A, Troeger C, et al. Morbidity, mortality and long-term consequences associated with diarrhoea from Cryptosporidium infection in children younger than five years: A meta-analysis. The Lancet Global Health, 2018;6(7): e758–e768. 10.1016/S2214-109X(18)30283-3

28. Schiaffino F, Colston J.M, Paredes Olortegui M, et al. The epidemiology and impact of persistent Campylobacter infections on childhood growth among children 0–24 months of age in resource-limited settings. EClinicalMedicine, 2024;76: 102841. 10.1016/j.eclinm.2024.102841

29. Gizaw Z, Yalew A.W, Bitew B.D, et al. Animal handling practices among rural households in north-west Ethiopia increase the risk of childhood diarrhoea and exposure to pathogens from animal sources. Environmental Health Insights, 2024;18: 11786302241245057. 10.1177/11786302241245057

30. Akpako B, Souley H, Ouattara A, et al. Qualitative molecular diagnostics may improve medical management of hospitalized severely malnourished children with diarrhoea: Analysis from Hôpital de l’Amitié Tchad–Chine, N’Djamena, Chad. 2016. Poster presented at DiDiMAS

31. Dickin S, Dagerskog L, Dione M, et al. Towards a One Health approach to WASH to tackle zoonotic disease and promote health and wellbeing. PLOS Water, 2025;4(5): e0000376. 10.1371/journal.pwat.0000376

32. Ruel M.T, Ashorn P, Berkley J.A, et al. Prevention of wasting and nutritional oedema: Evidence gaps identified during WHO guideline development. BMJ Global Health, 2025;10: e016314. 10.1136/bmjgh-2024-016314

33. World Health Organization. WHO guideline on the prevention and management of wasting and nutritional oedema (acute malnutrition) in infants and children under 5 years. Geneva: World Health Organization; 2023. ISBN: 978-92-4-008283-0.

34. De Hoop T, Molotsky A, Walcott R, et al. The role of nutrition-sensitive interventions in improving nutritional outcomes: findings from a systematic review and meta-analysis. Int J Equity Health, 2025; 24, 325. 10.1186/s12939-025-02596-y

35. Al Daccache M, Abi Zeid B, Hojeij L, et al. Systematic review on the impacts of agricultural interventions on food security and nutrition in complex humanitarian emergency settings. BMC Nutr, 2024;10, 60. 10.1186/s40795-024-00864-8

36. Sadler K, Kerven C, Calo M, et al. Milk matters: the role of livestock milk in pastoralist nutrition. Food and Nutrition Bulletin. 2012;33(2 Suppl):S66–S75. doi:10.1177/15648265120332S109

37. Muema J, Mutono N, Kisaka S, et al. The impact of livestock interventions on nutritional outcomes of children younger than 5 years old and women in Africa: a systematic review and meta-analysis. Frontiers in Nutrition. 2023;10:1166495. doi:10.3389/fnut.2023.1166495

38. Mutono N, Muema J, Bukania Z, et al. Effect of Behavioral Change Communication and Livestock Feed Intervention on Dietary Practices in a Kenyan Pastoral Community: A Randomized Controlled Trial. Nutrients. 2025; 17(18):2997. 10.3390/nu17182997

39. Kosek M.N, Haque R, Lima A.A, et al. Pathogen-specific burden of community diarrhoea in developing countries: A multisite birth cohort study (MAL-ED). The Lancet Global Health. 2020;8(6):e862–e873. doi:10.1016/S2214-109X(20)30174-5

40. George C.M, Oldja L, Biswas S, et al. Enteric infections, health outcomes, and economic burden in low-income settings: a review. The Lancet Infectious Diseases. 2022;22(4):e80–e92. doi:10.1016/S1473-3099(21)00376-1

41. Lin A, Arnold B.F, Afreen S, et al. Environmental enteric dysfunction indicators and child stunting: a multicountry pooled analysis. American Journal of Clinical Nutrition. 2021;114(3):1043–1051. doi:10.1093/ajcn/nqab128

42. Grace D, Mahoney R, Torr S, et al. One Health gains: lessons from integrated approaches to livestock and human disease control in low-resource settings. Philosophical Transactions of the Royal Society B. 2021;376(1821):20190701. doi:10.1098/rstb.2019.0701

43. Smith T.J, Mbale E, Zieff M.R, et al. Associations between maternal capabilities for care and nurturing care behaviours among mother-child dyads in Malawi and South Africa. PLOS Glob Public Health. 2025;5(9):e0005017

44. Waghode R.T, Yadav S.S, Ghooi R, et al. Investigating the association of maternal employment on the nutritional status of children up to 12 years: A systematic review. Nutrients. 2025;17(6):1059

45. Marshak A, Young H, Bontrager E.N, et al. The relationship between acute malnutrition, hygiene practices, water and livestock, and their programme implications in eastern Chad. Food and Nutrition Bulletin 2017;38(1): 117.

46. Venkat A, Marshak A, Young H, et al. Seasonality of Acute Malnutrition in African Drylands: Evidence From 15 Years of SMART Surveys. Food and Nutrition Bulletin. 2023;44(2_suppl):S94-S108. doi:10.1177/03795721231178344

## REFERENCES

1. International Labour Organization, Chad: Décret n° 11-055 PR/PM/MFPT du 21 janvier 2011 portant relèvement du Salaire Minimum Interprofessionnel Garanti (SMIG) et du Salaire Minimum Agricole Garanti (SMAG). 2011. N’Djamena, République du Tchad. [accessed 4 November 2025]. https://natlex.ilo.org/dyn/natlex2/natlex2/files/download/97322/TCD-97322.pdf

2. Akuoku J.K, Scott N, Manasseh Z, Moukénet A. Improving the allocative efficiency of nutrition investments in Chad: Results of Optima Nutrition analysis. 2023. [accessed 8 September 2025].

3. N’Diaye D.S, Wassonguema B, Nikièma V, et al. Economic evaluation of a reduced dosage of ready-to-use therapeutic foods to treat uncomplicated severe acute malnutrition in children aged 6–59 months in Burkina Faso. Maternal & Child Nutrition, 2021;17(3):e13118.

4. Isanaka S, Menzies N.A, Sayyad J, et al. Cost analysis of the treatment of severe acute malnutrition in West Africa. Maternal & Child Nutrition, 2017;13(4): e12398.

5. FAO. Preventing relapse of acute malnutrition: Impact assessment on locally produced supplementary food in northeastern Nigeria. 2024. Rome. 10.4060/cd1967en [accessed 20 October 2025]. https://fscluster.org/sites/default/files/documents/FAO_Northeastern%20Nigeria%20Preventing%20relapse%20of%20acute%20malnutrition%20Impact%20assessment%20on%20locally%20produced%20supplementary%20food%2C.pdf

6. Ritchie H. Half of all child deaths are linked to malnutrition. 2024. Published online at OurWorldinData.org. Retrieved from: https://ourworldindata.org/half-child-deaths-linked-malnutrition

7. Robinson L.A, Hammitt J.K, Cecchini M, et al. Reference case guidelines for benefit–cost analysis in global health and development. 2019. [accessed 20 April 2024] .https://www.ssrn.com/abstract=4015886

8. Gelli A, Kemp C.G, Margolies A, et al. Economic evaluation of an early childhood development center-based agriculture and nutrition intervention in Malawi. Food Security, 2022;14(1): 67–80. 10.1007/s12571-021-01203-6

9. Fidah M.F.A, Islam M.R, Amin R, et al. Cost of diarrhoea: A household perspective from seven countries in the Global Enteric Multicentre Study (GEMS). BMJ Paediatrics Open, 2025;9(1). [accessed 8 September 2025]. https://bmjpaedsopen.bmj.com/content/9/1/e003622

10. Usuf E. Mackenzie G, Sambou S, et al. The economic burden of childhood pneumococcal diseases in The Gambia. Cost Effectiveness and Resource Allocation, 2016;14: 4. 10.1186/s12962-016-0053-4

